# Domain mapping of disease mutations supports genetic testing of specific *SORL1* variants in familial Alzheimer’s Disease

**DOI:** 10.1101/2023.07.13.23292622

**Authors:** Henne Holstege, Matthijs W. J. de Waal, Niccolo’ Tesi, Sven J. van der Lee, Christa de Geus, Rosalina van Spaendonk, Maartje Vogel, Shahzad Ahmad, Najaf Amin, Philippe Amouyel, Gary W. Beecham, Céline Bellenguez, Claudine Berr, Joshua C. Bis, Anne Boland, Paola Bossù, Femke Bouwman, Jose Bras, Camille Charbonnier, Jordi Clarimon, Carlos Cruchaga, Antonio Daniele, Jean-François Dartigues, Stéphanie Debette, Jean-François Deleuze, Nicola Denning, Anita L. DeStefano, Oriol Dols-Icardo, Cornelia M. van Duijn, Lindsay A. Farrer, Maria Victoria Fernández, Wiesje M. van der Flier, Nick C. Fox, Daniela Galimberti, Emmanuelle Genin, Johan J.P. Gille, Benjamin Grenier-Boley, Detelina Grozeva, Yann Le Guen, Rita Guerreiro, Jonathan L. Haines, Clive Holmes, Holger Hummerich, M. Arfan Ikram, M. Kamran Ikram, Amit Kawalia, Robert Kraaij, Jean-Charles Lambert, Marc Lathrop, Afina W. Lemstra, Alberto Lleó, Richard M. Myers, Marcel M. A. M. Mannens, Rachel Marshall, Eden R. Martin, Carlo Masullo, Richard Mayeux, Simon Mead, Patrizia Mecocci, Alun Meggy, Merel O. Mol, Benedetta Nacmias, Adam C. Naj, Valerio Napolioni, J. Nicholas Cochran, Gaël Nicolas, Florence Pasquier, Pau Pastor, Margaret A. Pericak-Vance, Yolande A. L. Pijnenburg, Fabrizio Piras, Olivier Quenez, Alfredo Ramirez, Rachel Raybould, Richard Redon, Marcel J.T. Reinders, Anne-Claire Richard, Steffi G Riedel-Heller, Fernando Rivadeneira, Jeroen G. J. van Rooij, Stéphane Rousseau, Natalie S. Ryan, Pascual Sanchez-Juan, Gerard D. Schellenberg, Philip Scheltens, Jonathan M. Schott, Sudha Seshadri, Daoud Sie, Rebecca Sims, Erik A. Sistermans, Sandro Sorbi, John C. van Swieten, Betty Tijms, André G. Uitterlinden, Pieter Jelle Visser, Michael Wagner, David Wallon, Li-San Wang, Julie Williams, Jennifer S. Yokoyama, Aline Zarea, Marc Hulsman, Olav M. Andersen

**Affiliations:** Department of Human Genetics, Amsterdam UMC, location Vrije Universiteit Amsterdam, Amsterdam, The Netherlands; Alzheimer Center Amsterdam, Neurology, Amsterdam UMC, location Vrije Universiteit Amsterdam, Amsterdam, The Netherlands; Amsterdam Neuroscience, Neurodegeneration, Amsterdam, The Netherlands; Delft Bioinformatics Lab, Delft University of Technology, Delft, The Netherlands; Clinical Genetics, Human Genetics, Amsterdam UMC, Amsterdam, The Netherlands; Genome Analysis Laboratory, Human Genetics, Amsterdam UMC, location Vrije Universiteit Amsterdam, the Netherlands; Department of Epidemiology, Erasmus Medical Centre, Rotterdam, The Netherlands; LACDR, Leiden, The Netherlands; Nuffield Department of Population Health Oxford University; Univ. Lille, Inserm, CHU Lille, Institut Pasteur Lille, LabEx DISTALZ-U1167-RID-AGE - Facteurs de risque et déterminants moléculaires des maladies liées au vieillissement, Lille, France; The John P. Hussman Institute for Human Genomics, University of Miami, Miami, Florida, USA; Univ Montpellier, Inserm, INM (Institute for Neurosciences of Montpellier), Montpellier, France; Cardiovascular Health Research Unit, Department of Medicine, University of Washington, Seattle, WA, USA; Université Paris-Saclay, CEA, Centre National de Recherche en Génomique Humaine Evry, France; Experimental Neuro-psychobiology Laboratory, Department of Clinical and Behavioral Neurology, IRCCS Santa Lucia Foundation, Rome, Italy; Department of Neurodegenerative Science, Van Andel Institute, Grand Rapids, MI, USA; Division of Psychiatry and Behavioral Medicine, Michigan State University College of Human Medicine, Grand Rapids, MI, USA; Univ Rouen Normandie, Normandie Univ, Inserm U1245 and CHU Rouen, Departments of Genetics and CNRMAJ, F-76000 Rouen, France; Memory Unit, Neurology Department and Institut de Recerca Sant Pau, Hospital de la Santa Creu i Sant Pau, Universitat Autònoma de Barcelona, Sant Quintí 77-79, 08041, Barcelona, Spain; CIBERNED, Network Center for Biomedical Research in Neurodegenerative Diseases, National Institute of Health Carlos III, Madrid, Spain; Neurogenomics and Informatics Center, Washington University School of Medicine, St Louis, MO, USA; Psychiatry Department, Washington University School of Medicine, St Louis, MO, USA; Hope Center for Neurological Disorders, Washington University School of Medicine, St Louis, MO, USA; Department of Neuroscience, Catholic University of Sacred Heart, Fondazione Policlinico Universitario A. Gemelli IRCCS, Rome, Italy; University Bordeaux, Inserm, Bordeaux Population Health Research Center, France; Department of Neurology, Bordeaux University Hospital, Bordeaux, France; UKDRI at Cardiff, School of Medicine, Cardiff University, Cardiff, UK; Department of Biostatistics, Boston University School of Public Health, Boston, MA, USA; Framingham Heart Study, Framingham, MA, USA; Department of Neurology, Boston University School of Medicine, Boston, MA, USA; Department of Epidemiology, Boston University, Boston, MA, USA; Department of Medicine (Biomedical Genetics), Boston University, Boston, MA, USA; Dementia Research Centre, UCL Queen Square Institute of Neurology, London, UK; UK Dementia Research Institute at UCL, London, UK; Fondazione IRCCS Ca’ Granda, Ospedale Policlinico, Milan, Italy; University of Milan, Milan, Italy; Univ Brest, Inserm, EFS, CHU Brest, UMR 1078, GGB, F-29200, Brest, France; Division of Psychological Medicine and Clinical Neuroscience, School of Medicine, Cardiff University, Cardiff, UK; Quantitative Sciences Unit, Department of Medicine, Stanford University, Stanford, CA, USA; Department of Population and Quantitative Health Sciences, School of Medicine, Case Western Reserve University, Cleveland, Ohio, USA; Clinical and Experimental Science, Faculty of Medicine, University of Southampton, Southampton, UK; MRC Prion Unit at UCL, UCL Institute of Prion Diseases, London, UK; Division of Neurogenetics and Molecular Psychiatry, Department of Psychiatry and Psychotherapy, Faculty of Medicine and University Hospital Cologne, University of Cologne, Cologne Germany; Department of Internal Medicine, Erasmus Medical Centre, Rotterdam, The Netherlands; McGill University and Genome Quebec Innovation Centre, Montreal, QC, Canada; HudsonAlpha Institute for Biotechnology, Huntsville, AL, USA; Department of Human Genetics, Amsterdam UMC, University of Amsterdam, Amsterdam Reproduction and Development Research Institute Amsterdam, The Netherlands; Dr. John T. Macdonald Foundation Department of Human Genetics, University of Miami, Miami, Florida, USA; Institute of Neurology, Catholic University of the Sacred Heart, Rome, Italy; Taub Institute on Alzheimer’s Disease and the Aging Brain, Department of Neurology, Columbia University, New York, New York, USA; Gertrude H. Sergievsky Center, Columbia University, New York, NY, USA; Division of Gerontology and Geriatrics, Department of Medicine and Surgery, University of Perugia, Perugia, Italy; Division of Clinical Geriatrics, Department of Neurobiology, Care Sciences and Society, Karolinska Institutet, Stockholm, Sweden; Department of Clinical Genetics, Erasmus Medical Center, Rotterdam, The Netherlands; Department of Neuroscience, Psychology, Drug Research and Child Health University of Florence, Florence, Italy; IRCCS Fondazione Don Carlo Gnocchi, Florence, Italy; Penn Neurodegeneration Genomics Center, Department of Biostatistics, Epidemiology, and Informatics; University of Pennsylvania Perelman School of Medicine, Philadelphia, PA, USA; Penn Neurodegeneration Genomics Center, Department of Pathology and Laboratory Medicine, University of Pennsylvania Perelman School of Medicine, Philadelphia, PA, USA; Genomic And Molecular Epidemiology (GAME) Lab, School of Biosciences and Veterinary Medicine, University of Camerino (UNICAM) Camerino, 62032, Italy; Univ. Lille, Inserm, CHU Lille, UMR1172, Resources and Research Memory Center (MRRC) of Distalz, Licend, Lille France; Unit of Neurodegenerative diseases, Department of Neurology, University Hospital Germans Trias i Pujol, Badalona, Barcelona, Spain; The Germans Trias i Pujol Research Institute (IGTP), Badalona, Barcelona, Spain; Laboratory of Neuropsychiatry, Department of Clinical and Behavioral Neurology, IRCCS Santa Lucia Foundation, Rome, Italy; Department of Psychiatry and Glenn Biggs Institute for Alzheimer’s and Neurodegenerative Diseases, San Antonio, TX, USA; Department of Neurodegenerative Diseases and Geriatric Psychiatry, University Hospital Bonn, Medical Faculty, Bonn, Germany; German Center for Neurodegenerative Diseases (DZNE, Bonn), Bonn, Germany; Cluster of Excellence Cellular Stress Responses in Aging-Associated Diseases (CECAD), University of Cologne, Cologne, Germany; Université de Nantes, CHU Nantes, CNRS, INSERM, l’institut du thorax, Nantes, France; Institute of Social Medicine, Occupational Health and Public Health, University of Leipzig, Leipzig, Germany; Neurology Service, Marqués de Valdecilla University Hospital (University of Cantabria and IDIVAL), Santander, Spain; Amsterdam Reproduction & Development research institute, Amsterdam UMC, Amsterdam, The Netherlands; Univ Rouen Normandie, Normandie Univ, Inserm U1245 and CHU Rouen, Departments of Neurology and CNRMAJ, F-76000 Rouen, France; MRC UK Dementia Research Institute, Division of Psychological Medicine, Cardiff University, Cardiff, UK; Memory and Aging Center, Department of Neurology Weill Institute for Neurosciences, and Department of Radiology and Biomedical Imaging, University of California, San Francisco, CA, USA; Dept. of Biomedicine, Aarhus University, Denmark

**Keywords:** SORL1, SORLA, Alzheimer’s Disease, genetics, penetrance, age at onset, rare variants, disease risk, Alzforum Mutation Database

## Abstract

**Background:** Protein truncating variants (PTVs) in *SORL1* are observed almost exclusively in Alzheimer’s Disease (AD) cases, but the effect of rare *SORL1* missense variants is unclear.

**Methods:** To identify high-priority missense variants (HPVs), we applied ‘domain mapping of disease mutations’ on the 637 unique coding *SORL1* variants detected in 18,959 AD-cases and 21,893 non-demented controls.

**Results:** In this sample, PTVs and HPVs associated with respectively a 35- and 10-fold increased risk of early onset AD and 17- and 6-fold increased risk of overall AD. The median age at onset (AAO) of PTV- and HPV-carriers was 62 and 64 years, and *APOE*-genotype contributed to AAO-variability. The median AAO of PTV- and HPV-carriers is ∼8-10 years earlier than wild-type *SORL1* carriers, matched for *APOE*-genotype. Specific HPVs are highly penetrant and lead to earlier AAOs than PTVs, suggesting possible dominant negative effects.

**Conclusion:** Our results justify a debate on whether HPV carriers should be considered for clinical counseling.

## Background

The SORL1 protein, or SORLA, encoded by the *SORL1* gene, is the cargo-binding entity of the SORL1-retromer complex which regulates cargo-transport from the endosome back to the trans-Golgi network (‘retrograde’ pathway) and the transport of endocytosed receptors back to the cell surface (‘recycling’ pathway). Among SORL1’s numerous cargo is Amyloid-β and the amyloid precursor protein (APP): APP-binding by retromer-SORL1 accelerates APP-trafficking out of the early endosome, thereby warding off APP-cleavage and the subsequent formation and secretion of Amyloid-β(1), which links impaired SORL1 with hallmark processes of Alzheimer’s disease (AD). Genetic variants in SORL1 have been linked to AD risk since 2007(2). Genome wide association studies (GWAS) reported that common single nucleotide polymorphisms (SNPs) in or near *SORL1* associated with AD, though with limited effects(3–6). Furthermore, large case-control sequencing studies reported that rare coding variants in *SORL1* had a considerable effect on AD(7–11).

SORL1 is a multidomain, 2,214 amino-acid protein, encoded by 48 exons, such that by virtue of its size, *SORL1* genetic sequence is vulnerable for acquiring mutations. More than 3,000 coding variants are listed in GnomAD(12), with effects ranging from negligible to deleterious on protein function. Potentially damaging *SORL1* variants affect as many as 2.75% of all genetically unrelated early onset AD cases (EOAD, with Age at Onset (AAO) <65 years) and 1.5% of genetically unrelated late onset AD cases (LOAD, with AAO >65 years)(13). Of these, protein truncating variants (PTV) occur almost exclusively in AD cases(10, 13), suggesting that *SORL1*’s haploinsufficiency is highly penetrant and may be causative of AD(7, 10, 13, 14). Indeed, a recent report described a Peruvian pedigree affected with a p.Trp1673Ter PTV with an inheritance pattern of AD suggestive of autosomal dominant AD (ADAD)(15). However, most *SORL1* variants observed in AD cases are *rare missense SORL1* variants, mostly unique to one person and their family-members, some may increase risk or cause disease, while many are benign(16–18).

In our clinics, we have identified several large families affected with (early onset) AD with an inheritance pattern suggestive of ADAD, in which the proband carries a *SORL1* variant. However, since DNA of affected and unaffected relatives is commonly unavailable for segregation analyses, it remains unclear whether the *SORL1* variant is the cause of the ADAD in these families(18). Over the past years, several (assembled) pedigrees were reported that suggest high penetrance of specific *SORL1* variants(8, 14, 17–21). However, in contrast to certain variants in the classic *PSEN1/PSEN2/APP* ADAD genes(22), no fully penetrant *SORL1* variants have been identified. While incomplete penetrance has been described in ADAD genes as well(23, 24), this lack of proper AD risk estimates associated with the diverse *SORL1* variants causes uncertainty of their clinical relevance. Consequently, even protein truncating variants in *SORL1* are classified as ‘VUS’: **<u>v</u>**ariant of **<u>u</u>**nknown **s**ignificance (ClinVar(25)) and such variants are not routinely communicated to patients, despite accumulating evidence that specific *SORL1* variants are highly penetrant(14). For clinical geneticists to consider including *SORL1* variants in AD diagnostics a better understanding of *SORL1* variant pathogenicity and their effects on AAO is imperative.

Here, we took advantage of the increasing knowledge of SORL1 function (**Fig 1**) and we learned from the effect of specific missense variants on the function of proteins that share domains with SORL1 (accompanying manuscript). This manual domain-mapping of disease mutations (DMDM) approach led to the prioritization of functionally relevant *SORL1* variants. We applied this prioritization strategy to *SORL1* variants identified in sequencing data of 18,959 AD cases and 21,893 non-demented controls(13) and identified high-priority (HPV), moderate-priority (MPV), low-priority (LPV) and no-priority (NPV) missense variants. We investigated the AD risk and AAO of AD associated with variants from these groups, and further categorized high-priority missense variants into specific subcategories. With this work, we aim to provide insight into *SORL1* variant pathogenicity, and effects on AAO. With this, we aim to ultimately contribute to the discussion whether identifying and disclosing SORL1 variant in AD patients is beneficial (26, 27).

**Figure 1:**
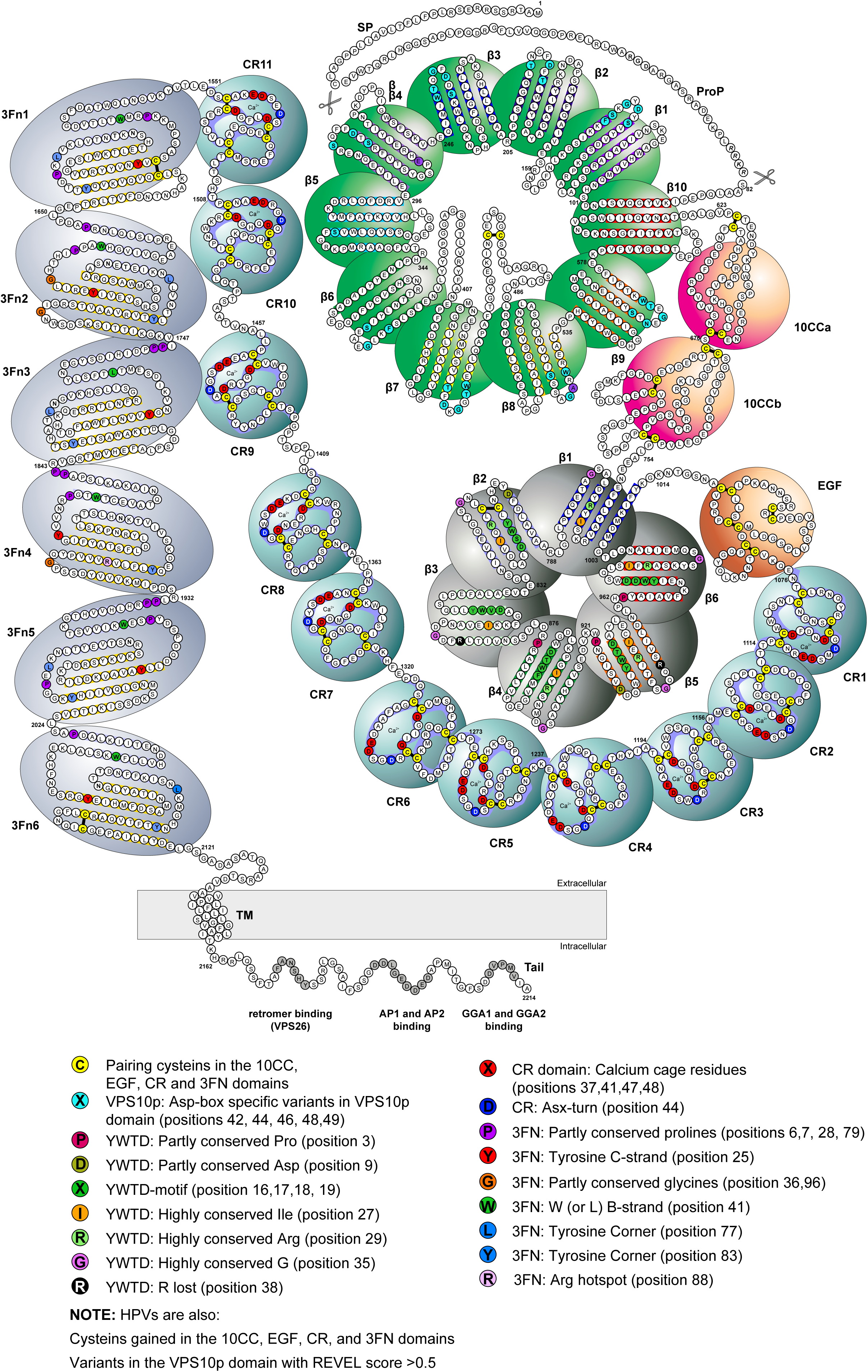
Schematic of the SORL1 domains and their function, highlighting the variants with the strongest effects on AD risk, that may exist in several domain-repeats. SORL1 is a large, 2,214 amino acid multi-domain protein, which includes multiple repeated domain elements, each of which includes many strictly or moderately conserved residues important for protein domain folding and/or the binding of ligands. When the SORLA protein is transcribed at the ribosome, the protein signal peptide (res 1-28) is cleaved off upon translocation to the endoplasmatic reticulum (ER). During its transport through the trans-Golgi-Network, the SORL1 protein undergoes several post-translational modifications, including N- and O-glycosylation at multiple sites(59, 60). During this maturation process, the pro-domain (res 29-81) is speculated to prevent binding of certain ligands to the VPS10p-domain in the endoplasmatic reticulum (ER), where receptor and ligand are co-expressed. The pro-domain is cleaved off by Furin once SORL1 leaves the trans-Golgi-Network, where it can engage in ligand binding and trafficking. The VPS10p-domain (res 82-617), a ten-bladed β-propeller domain, is a flat disc that is stabilized at its bottom face by the 10CC-domain (res 618-753). At its top face, the VPS10p-domain binds ligand, it further has a large hydrophobic tunnel at its center, allowing interaction with small lipophilic ligands such as the Amyloid-β peptide. The domain contains two protrusions (loop structures, loop L1 and loop L2): the VPS10p-domain can bind ligand at neutral pH and while L1 blocks part of the tunnel, the L2 protrusion pushes the ligand against the tunnel wall. After trafficking to a more acidic part of the cell (i.e. the lysosome), L1 and L2 change conformation and release the ligands from the VPS10p pore. C-terminal to the 10CC-domain is a ligand-binding YWTD β-propeller (res 754-1013), which is stabilized at its bottom face by an EGF-domain (res 1014-1074, fully encoded by exon 22), such that ligand-interactions with both the VPS10p and YWTD β-propellers occur at their top faces. The combined action of VPS10p β-propeller and the YWTD β-propeller might enable interactions with large ligands including co-receptors in multimeric complexes, or large soluble ligands requiring two adjacent β-propellers for efficient binding, akin to what was recently identified for LRP4/Agrin/MusK signalling complex(61). C-terminal to the EGF-domain comes the CR-cluster (res 1075-1550) which is the interacting site of at least half of the SORL1-ligands, including APP. This cluster is like a flexible necklace composed of 11 unique ∼40 amino-acid CR-domains, each encoded by a single exon (exons 23-33), that each form the ‘pearls’ on the string. These can wrap around larger ligands and engage in minimal motif interactions with multiple sites of a ligand, leading to high-affinity ligand binding. Each CR-domain includes 16 strictly conserved amino acids, including six disulfide bridge-forming cysteines, such that all CR-domains have a similar compact folding. Each CR-domain further contains four conserved residues that form an octahedral ‘calcium-cage’ which stabilizes the domain, and in combination with two backbone carbonyls, coordinates a calcium ion, which is critical for calcium-dependent domain folding. The side chains of these two residues engage in minimal-motif ligand binding, which explains why ligand binding to CR-domains relies on Ca^2+^. Substituting these may impair the binding of specific ligands, but do not affect overall folding and stability of CR-domains(62). Preliminary evidence suggests that perturbation of the calcium-cage on the other hand, may lead to a misfolded SORL1 protein that is retained in the ER(63). C-terminal of the CR-cluster is the 3Fn-cassette (res 1551-2121), containing 6 ellipsoid 3FN-domains, each containing several conserved and partly conserved residues, and involved in SORL1 dimerization(46). Therefore, genetic variants affecting one of the conserved residues in 3FN-domain is likely to disturb SORL1 dimerization(19). Lastly, SORL1 has a transmembrane and cytoplasmic tail domain (res 2122-2214) which can interact with the VPS26 subunit of the retromer complex(64). Recent evidence suggests that SORL1 matures (by *N-* and *O-*glycosylation) at the ER/Golgi in a monomer form, then travels to the endosome where it dimerizes at its 3FN-domain and its VPS10-domain(19). The dimerized SORL1 uses its cytoplasmic tail domain to interact with the VPS26 subunit of the retromer complex, allowing SORL1 to engage in retromer-dependent cargo trafficking through the endolysosomal system.

## Methods

### Samples

We extracted *SORL1* genetic variants from the assembled whole exome sequencing (WES) and whole genome sequencing (WGS) data as previously described(13), which includes data contributed by the European ADES cohort and the ADSP, StEP-AD, Knight-ADRC and UCSF/NYGC/UAB cohorts, Procedures for AD diagnosis were described previously(13), and occurred according to the National Institute of Neurological and Communicative Disorders and Stroke-Alzheimer’s Disease and Related Disorders Association criteria(28) or the National Institute on Aging-Alzheimer’s Association criteria(29). Carriers of a pathogenic variant(s) in *PSEN1*, *PSEN2*, or *APP* or in any other gene associated with Mendelian dementia were excluded. Relatives up to 3^rd^ degree of relatedness were excluded to avoid any family-based effects(30).

Variant carriers were identified by determining posterior genotype dosages, using genotype probabilities and the frequency of the variant in the dataset as a prior, allowing us to take genotyping uncertainty into account (previously described in Methods and Supplementary Note of Holstege and Hulsman et al., 2022(13)). For analysis, we considered variants that had at least 1 carrier, that is, at least one sample with a posterior dosage >0.5. Application of our comprehensive quality control procedures allowed the retention of more AD cases and controls for a *SORL1*-specific analysis, compared to our previously published genome-wide analysis(13). We further increased analysis power by including *SORL1* variants identified in individuals with non-European ancestry, the rationale for this is that rare *SORL1* variants have been reported to associated with AD risk across all populations studied thus far(31). To correct our analysis for population-specific effects, we performed a principal component analysis (PCA) including the first 6 principal components as covariates in our analysis. Together, after quality control (QC), the current sample included *SORL1* sequences from 40,852 individuals: 18,959 AD cases and 21,893 non-demented controls, comprising 12.9% African, 0.06% East Asian, 0.61% South Asian, 5.84% Admixed Americans and 80.59% European: for a PCA representing the sample by population background see **Figure S1**.

### Variant Quality Control

The raw sequence data was processed with a uniform pipeline as described previously(13). In brief, the data was processed relative to the GRCh37 reference genome, after which extensive quality control was applied which led to the exclusion of likely false positive variant-calls from analysis. Other variants were excluded due to differential missingness, positions for which coverage across cases and controls differed >5%. These included all variants in exon 1 (res 1-95), which codes for the signal peptide (res 1-28), the pro-domain (29-81), and the first 10 residues of the VPS10p-domain. See **Table S3** for excluded variants.

### Variant annotation

We annotated *SORL1* variants that occur in the canonical transcript (T260197). All variants were annotated with the ‘non-neuro popmax’ minor allele frequency (MAF) using the GnomAD database version v.2.1.1. Variants that were absent from GnomAD database were annotated by their MAF in the total current sample. Variants with MAF>0.05% (which relates to having at least 21 carriers in this sample) were considered less-rare. Variants with a MAF<0.05% were considered rare, and included in a domain-specific rare variant burden analysis (**Fig 2**).

**Figure 2:**
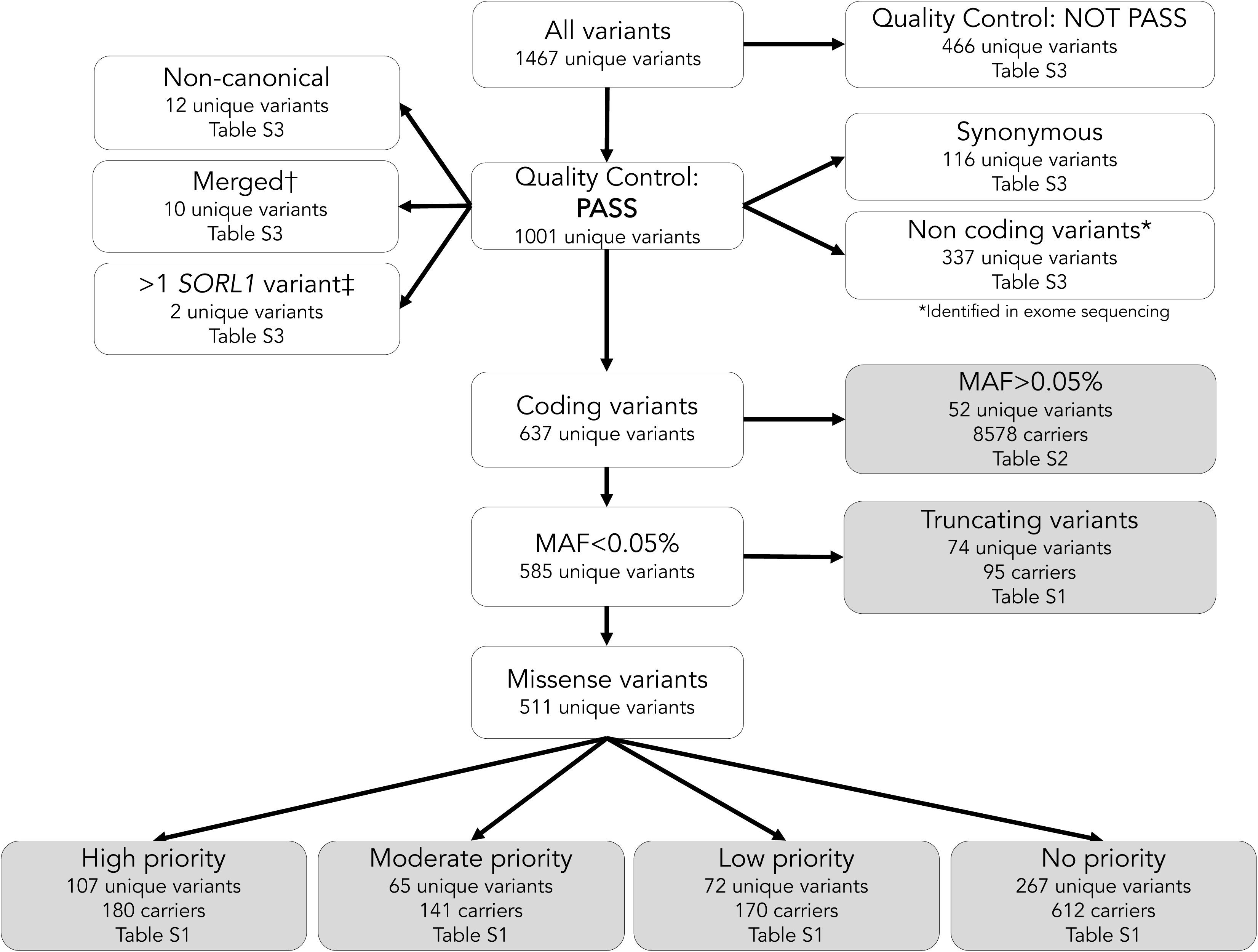
Flowchart of selection procedures for *SORL1* variant subtypes. *Non-coding variants were identified in the padding of exome sequencing or in exome excerpts from whole genome sequences. †Variants were merged into one when they were juxtaposed and in cis. ‡The 27 individuals who carried more than one rare *SORL1* variant were grouped according to the variant with the highest priority. Among these was one case (AAO 46, *APOE-ε3*/*ε4*) who carried a p.C1453F ONC variant in combination with a PTV, and one case (AAO 55, *APOE-ε3*/*ε4*) carried a 11:121459965:T>TG splice variant in combination with a PTV; both were grouped with PTV carriers.

We used the Variant Effect Predictor in Ensembl database (VEP, version v.94.542) to identify variants with a possible consequence on protein function. Missense variants were annotated with the rare exome variant ensemble learner (REVEL) score(32), which ranges from 0 (no predicted effect on protein function), to 1 (high predicted effect). Variants comprising two consecutive missense variants that give rise to two consecutive amino acid substitutions could not be annotated by REVEL and were conservatively annotated according to the substitution with the lowest REVEL score. PTVs were identified using the Loss-Of-Function Transcript Effect Estimator (LOFTEE, version v.1.0.2)(33), which annotates nonsense, frameshift and splice variants that lead to protein truncation as 1, and non-PTVs as 0. Note that PTVs in the last exon 48 should be considered deleterious, as this encodes the cytoplasmic tail domain which includes the FANSHY motif (for retromer binding), DDLGEDDED motif (sequence for binding cytoplasmic AP1 and AP2) and the DVPMV motif (sequence for GGA1 and GGA2 binding) which are all necessary for cellular trafficking and activity. Since LOFTEE did not annotate exon 48 PTVs, we manually included them in the PTV list. Splice variants with LOFTEE score 0 were evaluated using Splice AI(34) and those with a potential splice effect were evaluated manually by a trained clinical geneticist (MV), and those with expected effects on splicing were added to the list of PTVs.

### Prioritization of rare missense variants

Rare missense variants (MAF <0.05%) were separated into high-priority variants (HPVs), moderate-priority variants (MPVs) low-priority variants (LPVs) and no-priority variants (NPVs), according to the variant prioritization scheme developed by Andersen *et al*. (accompanying manuscript(35)), and presented in **Table 2**, identified variants are listed in **Table S1**. HPVs were identified as according the DMDM analysis(35), independent of REVEL score, and are indicated in **Fig 3** and listed in **Table S4** An exception is the p.VPS10p-domain and 10CC-domain combination, as the 5 members of the VPS10p-receptor protein family hold no/only few disease-associated variants. Therefore, DMDM analysis was not possible for the variants in the VPS10p-domain, such that, apart from the variants involving cysteines in VPS10p loops L1 and L2, we relied on applying a REVEL score threshold of >0.5, for which we previously found the strongest effect on AD risk(13). MPVs are rare missense variants annotated as moderate-priority by DMDM as indicated in **Fig 3** and listed in **Table S5**). LPVs are rare missense variants with a REVEL score >0.5 that are not in the VPS10p-domain, and NPVs comprise all remaining rare variants that are not HPV, MPV or LPV.

**Figure 3:**
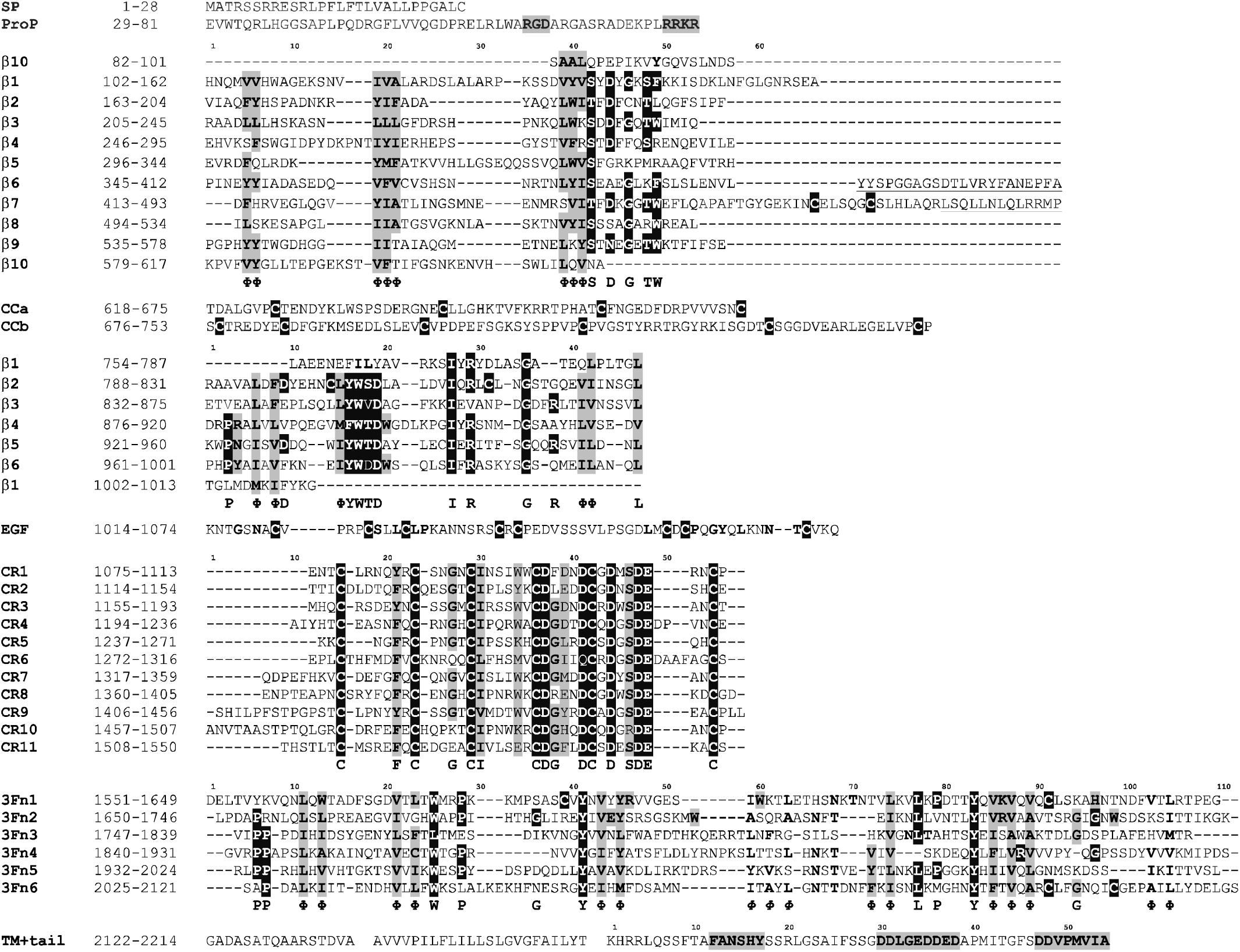
Protein sequence alignments per SORL1 subdomain. The protein sequence was aligned for each repeat in each SORL1 subdomain, revealing conserved residues. Residues likely to harbor deleterious mutations based on either domain sequence conservation or because the DMDM analysis were prioritized. **Black:** High-priority variants (HPVs) affect residues annotated in black; **Grey:** Moderate-priority variants (MPVs) affect residues indicated in grey.

### Rare variant association with Alzheimer’s Disease

Effects for less-rare variants (MAF>0.05%) on AD were evaluated per variant using a logistic regression model (**Table S2)**, while correcting for population effects using PCA components (PC1-PC6) (**Fig S1**). Rare variants with a MAF<0.05% were considered in a domain-specific rare variant burden analysis (**Fig 2**). We associated carrying a variant with MAF <0.05% appertaining to a specific priority category with AD (burden test). We repeated analyses stratifying EOAD cases (AD-aao <65 years) and LOAD cases (AD-aao >65 years), relative to the same group of controls. Since the number carriers of specific variants was low, the significance of the association was determined using a Fisher exact test, and corrected for multiple testing (Bonferroni), p_adj_<0.05 was considered significant. Calculations were performed using the epiR package (v.2.0.38).

### Age at onset curves

Since controls were overall much younger than cases, we compared the effect of different *SORL1* variants on AAO variants in a case-only AAO analysis. We used Kaplan-Meier survival analysis (CI of 95%) to estimate AAO curves (in R using Survival (v.3.3-1). For each variant priority category, we compared the median AAO with the median AAO of *SORL1*-WT carriers in our cohort. Log rank tests were performed to test for differences between AAO curves. Additionally, we stratified according to *APOE* genotype.

### Effect of APOE

We investigated a possible interaction-effect between *APOE* genotype (no, one or two APOE-*ε4* alleles) and *SORL1* priority category. To avoid confounding an interaction signal by samples in which APOE status was part of selection criteria, we performed an interaction analysis on ADES dataset only, for which there was no selection for *APOE*-genotype. We tested for both additive effect (*SORL1*+*APOE*) and interactive effect (*SORL1*\**APOE*) using a logistic regression models adjusted for PCA components (PC1-PC6). Interaction effects were tested using a Likelihood Ratio Test.

## Results

After quality control we observed 637 unique coding *SORL1* variants across 18,959 genetically unrelated cases (mean age: 72.4±10.7, 59.5% females, 50.4% APOE-ε4 carriers) and 21,893 controls (mean age: 71.1±16.7, 58.23% females, 17.2% APOE-ε4 carriers) (**Table 1**). These included 585 variants with MAF<0.05%: 74 PTVs and 511 missense variants that were further stratified into 267 NPVs, 72 LPVs, 65 MPVs, and 107 HPVs, (see Online Methods, **Table S1**, **Fig 2)**, and 52 variants with MAF>0.05% (**Table S2**).

**Table 1.**
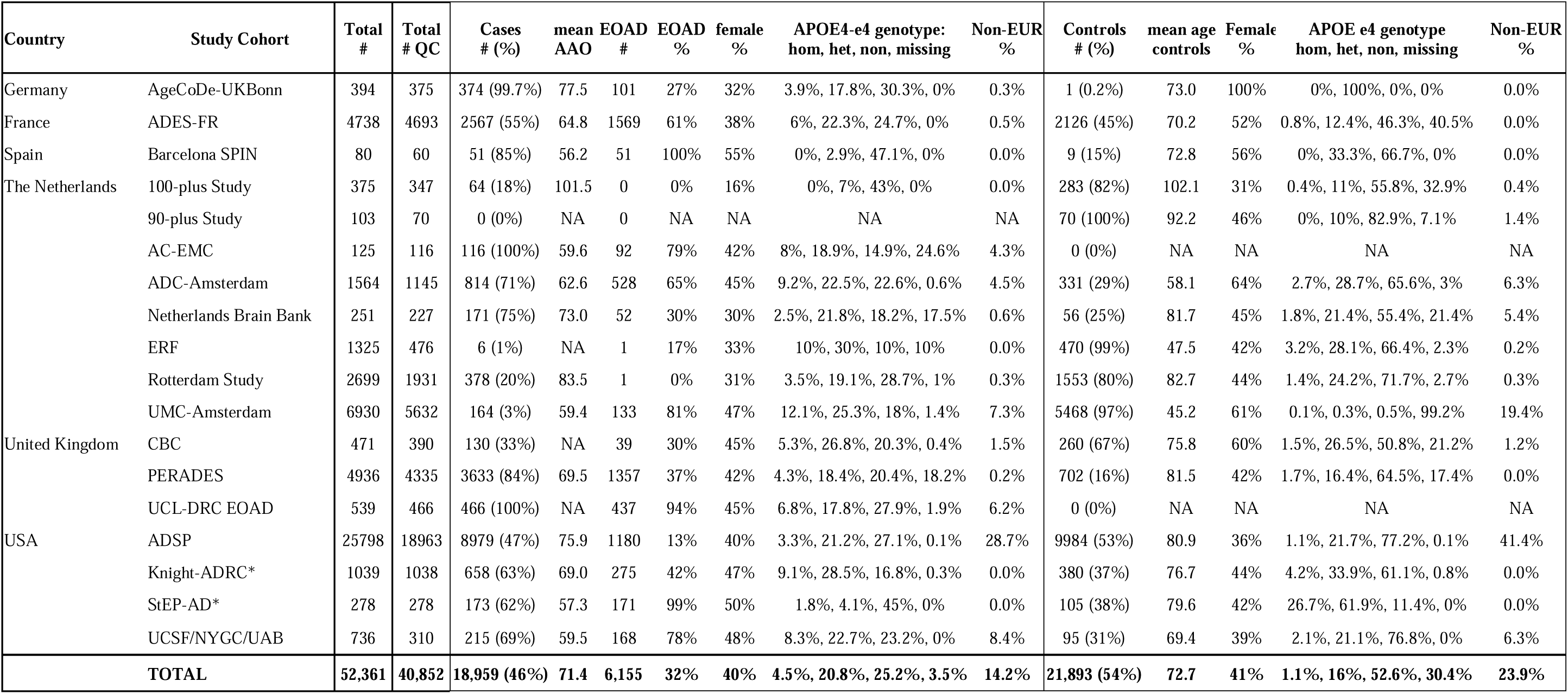
Cohort characteristics: Characteristics of the samples contributed by each study, grouped by country. Sequencing is based on exome sequencing, except for UCS/NYG/UAB, which was based on whole genome sequencing (WGS) data. Respectively 11% and 10% of the ADSP AD cases and controls comprised WGS data, and 30% of the ADES-FR cases. QC: quality control: next to selection based on technical procedures as described in the Online Methods, we included only genetically unrelated individuals (Identity By Descent >3^rd^ degree relations). A.A.O: mean age at onset; A.L.S. mean age at last screening. EOAD: early onset cases, a.a.o. ≤65. Ages annotated “>89” were set to 90. See supplement for detailed cohort descriptions. Non-Europeans: Africans (AFR), Admixed Americans (AMR), East Asicans (EAS), and South Asians (SAS). *Samples from Knight-ADRC and StEP-AD cohorts were extracts of the coding sequences of the *SORL1*, *TREM2*, *ABCA7*, *ATP8B4*, *ABCA1*, *ADAM10*, *RIN3*, *CLU*, *ZCWPW1*, *ACE, and CBX3* genes as described previously(13), and based on a separate PCA on these extracts, samples were annotated as EUR as we did not identify population outliers based on these exome extracts. To distinguish non-Europeans from Europeans (Table 1) we trained a k-nearest neighbor classifier on the first 10 PCA components, using the 1000G samples (SKLearn 26 v0.20.3, k=10).

**Table 2.** Prioritization scheme of rare variants. Rare variants with MAF<0.05% were considered for prioritization. HPV and MPVs affect residues corresponding to the black and grey residues in **Figure 3**. **^1^MAF**: Variant Minor Allele Frequency in the non-neuro pop-max dataset (Gnomad v.2.1.1). When unavailable, we used the variant MAF in the sample. Non-neuro sample in GnomAD is the sample without individuals with neurological diseases. The pop-max is the population with the highest frequency (pop-max). ^2^REVEL (Ioannidis et al): Variant effect prediction algorithm: Rare Exome Variant Ensemble Learner. Scores range from 0 to 1 and variants with higher scores are predicted to be more deleterious.

| Prioritization category | Selection criteria |
| --- | --- |
| PTV: protein truncating variants | All truncating variants: i.e. nonsense, frameshift and splice variants* |
| HPV: High-priority missense variants | Variants that affect *high-priority* residues (**Table S4**)<br>Variants in the p.VPS10p and 10CC-domains *and* REVEL score $\geq 0.50$ |
| MPV: Moderate-priority variants | Variants that affect *moderate-priority* residues (**Table S5**) |
| LPV: Low-priority variants | Variants *not prioritized* with REVEL score $\geq 0.50$ |
| NPV: No-priority variants | All remaining variants |

### PTV, Protein Truncating Variants

The 74 PTV variants (nonsense, frameshift and splice variants), were observed in 89 cases and 6 controls (<50 [n=1], 50–75 [n=3], >75 [n=2] at last screening) and associated with an overall 17.2-fold increased risk of AD (95%CI 7.5–39.3; p =1.2x10^-21^). PTVs associated with a 35.3-fold increased risk of EOAD (95%CI 15.2–81.8; p =6.2x10^-31^) and an 8.6-fold increased risk of LOAD (95%CI 3.6–20.6; p=3.8x10^-7^) (**Table 3**). A survival analysis indicated that the median AAO of a *SORL1*-PTV carrier was 62 years (10%-90% range: 52-78), 10-years earlier (95%CI -12– -8; 2.66x10^-11^) than the median AAO of *SORL1*-wild-type (WT) carriers in our sample at 72 (10%-90% range: 56–87) (**Fig 4A**, **Table 3**).

**Figure 4.**
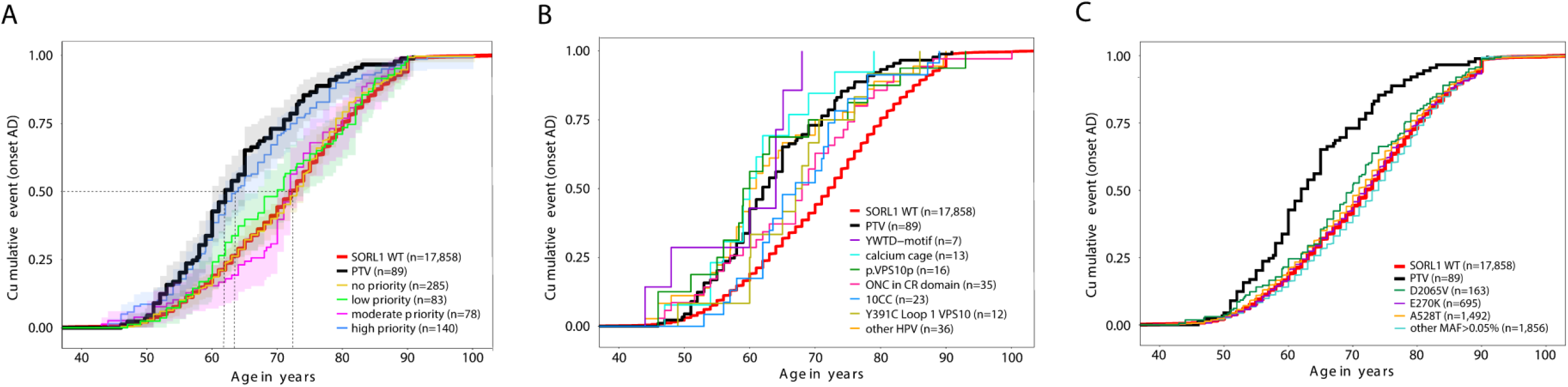
Age at onset analysis of case-carriers. AD cases were categorized according to the variant subtype they carry. **A)** Age at onset analysis for carriers of missense variants annotated as high-, moderate-, low- and no-priority, compared to carriers of PTVs and SORL1 WT carriers. **B)** Age at onset analysis of carriers of specific high-priority missense variants, relative to carriers of PTVs and WTs. **C)** Age at onset analysis of less rare variants (MAF>0.05%). Horizontal lines indicate the age at which 50% of all carriers with the same variant category have AD. Differences in ages at onset between carriers of variants appertaining to specific variant-groups are shown in **Table S6.**

**Table 3.** Effect of variant subtypes on AD risk, and effect on age at onset: AAO: Age at onset; OR: odds ratio, calculated using a Fisher’s Exact test. Note that all effect sizes were calculated relative to the same control group, which was relatively young, such that effect sizes may be conservative. *Odds rations for all variants with MAF>0.05% were calculated using a logistic regression model on the minor allele dosages. P values were corrected for multiple testing using Bonferroni. When common enough, variants were also imputed in the latest GWAS(37) (**Table S2**), association statistics for the most common variants were: E270K: OR= 1.02 95%CI 0.95-1.09, p = 5.89E-01; A528T OR=1.11, 95%CI 1.07-1.15, p value =5.79E-08. For the D2065V variant we observed an OR=1.36 (95%CI 1.2-1.54) with a p value=1.61E-06, which is in the opposite direction compared to the AD association observed in the current exome sequencing dataset (OR=0.87, p=0.18). Assuming no inaccuracies in the variant imputation or variant annotation, this difference indicates the limited power of the smaller exome sample size to correctly reflect association statistics, hence the meaningless p-value. Possibly, effects may be weaker in EOAD cases, which are more prevalent in the exome study compared to the GWAS study.

| Coding rare variants | Variant Subcategory | carriers | # unique variants | Variant effect on AD risk |  |  |  |  |  | Effect on AAO |  |  |
| --- | --- | --- | --- | --- | --- | --- | --- | --- | --- | --- | --- | --- |
|  | domain residues | all/EOAD/LOAD/ controls | all/EOAD/LOAD/controls | All carriers OR | All carriers p value | EOAD OR | EOAD p value | LOAD OR | LOAD p value | Median AAO (10%-90% IPR) | Δ AAO vs WT (95% CI) | p value* |
| **SORL1 WT** |  |  |  |  |  |  |  |  |  |  |  |  |
| WT |  | 31,324/4,642/9,677/17,005 | N/A | 0.89 (0.85 - 0.93) | 8.68E-06 | 0.88 (0.83 - 0.94) | 6.2E-3 | 0.89 (0.85 - 0.94) | 2.3E-04 | 72 (56 - 87) | NA | N/A |
| **Non-rare variants** |  |  |  |  |  |  |  |  |  |  |  |  |
| —MAF>0.05%* | E270K* | 1,409/212/479/718 | 1/1/1/1 | 1.02 (0.92 - 1.13) | 6.88E-01 | 0.94 (0.81 - 1.1) | 4.3E-01 | 1.1 (0.95 - 1.2) | 2.9E-01 | 72 (56 - 87) | 0 (-1 — 0) | 1 |
|  | A528T* | 2,719/495/961/1,263 | 1/1/1/1 | 1.2 (1.08 - 1.26) | 6.15E-05 | 1.1 (1.03 - 1.27) | 1.2E-02 | 1.2 (1.08 - 1.28) | 1.6E-04 | 71 (55 - 86) | -1 (-2 — -1) | 1.7E-01 |
|  | D2065V* | 353/55/106/192 | 1/1/1/1 | 0.9 (0.7 - 1.07) | 1.79E-01 | 0.87 (0.64 - 1.18) | 3.7E-01 | 0.9 (0.68 - 1.09) | 2.2E-01 | 70 (53.1 - 85) | -2 (-4 — -1) | 7.1E-01 |
|  | All other | 4,035/500/1,314/2,221 | 48/34/38/45 | 1.0 (0.95 - 1.09) | 6.0E-01 | 1.0 (0.91 — 1.12) | 8.2E-01 | 1.0 (0.95 - 1.10) | 6.4E-01 | 74 (57 - 87) | 2 (-1 — 1) | 1 |
| **Protein Truncating Variants** |  |  |  |  |  |  |  |  |  |  |  |  |
| PTV | Total | 95/59/30/6 | 74/52/25/6 | 17.2 (7.5 - 39.3) | 1.2E-21 | 35.3 (15.2 - 81.8) | 6.2E-31 | 8.6 (3.6 - 20.6) | 3.8E-07 | 62 (52 - 78) | -10 (-12 — -8) | 2.7E-11 |
| **Rare Missense variants** |  |  |  |  |  |  |  |  |  |  |  |  |
| HPV: high priority | Total | 180/80/71/29 | 107/62/50/23 | 6.1 (4.1 - 9.0) | 5.3E-24 | 9.9 (6.5 - 15.2) | 7.8E-29 | 4.2 (2.7 - 6.5) | 1.3E-10 | 64 (53 - 79) | -8 (-10 — -6) | 5.3E-09 |
| —VPS10p | Total | 43/22/16/5 | 27/17/10/5 | 8.8 (3.5 - 22.3) | 3.4E-07 | 15.7 (5.9 - 41.5) | 2.2E-09 | 5.5 (2.0 - 15) | 9.5E-03 |  |  |  |
|  | —Cysteins gained & cysteins lost L1/L2 | 18/8/8/2 | 6/4/2/2 | 9.2 (2.1 - 40.2) | 6.7E-03 | 14.2 (3.0 - 67.1) | 4.5E-03 | 6.8 (1.5 - 32.2) | 1.9E-01 |  |  |  |
|  | —Asp-box, REVEL>0.5 | 6/3/3/0 | 5/3/3/0 | NA | 2.8E-01 | NA | 3.0E-01 | NA | 1 |  |  |  |
|  | —Remaining p.VPS10p variants, REVEL>0.5 | 19/11/5/3 | 16/10/5/3 | 6.2 (1.8 - 21.2) | 2.6E-02 | 13.1 (3.6 - 46.8) | 2.8E-04 | 2.9 (0.7 - 11.9) | 1 | 59.5 (46 - 83) | -12.5 (-16 — -5) | 1 |
| —10CC | Total | 30/11/13/6 | 19/8/10/3 | 4.6 (1.9 - 11.3) | 8.7E-03 | 6.5 (2.4 - 17.7) | 5.0E-03 | 3.7 (1.4 - 9.8) | 1.9E-01 | 67 (57 - 78) | -5 (-9 — 0) | 1 |
| —YWTD | Total | 25/13/7/5 | 12/9/6/5 | 4.6 (1.7 - 12.3) | 2.6E-02 | 9.3 (3.3 - 26.0) | 2.1E-04 | 2.4 (0.8 - 7.5) | 1 |  |  |  |
|  | —YWTD-motif (17, 18, 19, 20) | 8/6/2/0 | 6/5/2/0 | NA | 6.2E-02 | NA | 3.1E-03 | NA | 1 | 64 (44 - 68) | -8 (-24 — NA) | 2.5E-03 |
|  | —Highly conserved residues (29, 35) | 8/2/3/3 | 3/2/2/3 | 1.9 (0.5 - 8.1) | 1 | 2.374 (0.4 - 14.2) | 1 | 1.7 (0.4 - 8.5) | 1 |  |  |  |
|  | —partly conserved residues (9, 38) | 8/4/2/2 | 2/1/2/2 | 3.5 (0.7 - 17.2) | 1 | 7.1 (1.3 - 38.9) | 6.7E-01 | 1.71 (0.2 - 12.1) | 1 |  |  |  |
| —EGF | Cysteins gained or lost | 2/1/0/1 | 2/1/0/1 | 1.15 (0.1 - 18.5) | 1 | 3.6 (0.2 - 56.9) | 1 | NA | 1 |  |  |  |
| —CR | Total | 57/23/26/8 | 36/21/19/6 | 7.1 (3.4 - 15.0) | 1.9E-08 | 10.3 (4.6 - 23.0) | 2.3E-08 | 5.6 (2.5 - 12.3) | 8.6E-05 |  |  |  |
|  | —Calcium Cages (D, D, D, E) 37, 41, 47, 48 | 13/9/4/0 | 12/9/4/0 | NA | 1.3E-03 | NA | 3.4E-05 | NA | 5.1E-01 | 60 (54 - 73) | -12 (-16 — NA) | 7.7E-04 |
|  | —Cysteins gained or lost | 42/12/22/8 | 22/10/15/6 | 4.9 (2.3 - 10.6) | 1.8E-04 | 5.3 (2.2 - 13.1) | 7.3E-03 | 4.7 (2.1 - 11.0) | 2.0E-03 | 68 (53 - 82) | -4 (-9 — 1.3) | 1 |
|  | —Asx-turn (D) (44) | 2/2/0/0 | 2/2/0/0 | NA | 1 | NA | 1 | NA | NA |  |  |  |
| —3Fn | Total | 15/8/5/2 | 10/5/4/2 | 7.5 (1.7 - 33.3) | 7.8E-02 | 14.2 (3.0 - 67.1) | 4.5E-03 | 4.3 (0.8 - 22.0) | 1 |  |  |  |
|  | —Partly conserved glycines (36, 96) | 2/1/1/0 | 2/1/1/0 | NA | 1 | NA | 1 | NA | 1 |  |  |  |
|  | —Partly conserved prolines (6, 7, 79) | 4/2/2/0 | 4/2/2/0 | NA | 1 | NA | 1 | NA | 1 |  |  |  |
|  | —highly conserved residues (25, 41, 77, 83) | 9/5/2/2 | 4/2/1/2 | 4.0 (0.8 - 19.5) | 1 | 8.9 (1.7 - 45.9) | 2.0E-01 | 1.7 (0.2 - 12.1) | 1 |  |  |  |
| MPV: Moderate priority | all | 141/20/60/61 | 65/16/39/33 | 1.5 (1.1 - 2.1) | 3.9E-01 | 1.2 (0.7 - 1.9) | 1 | 1.7 (1.2 - 2.4) | 1.2E-01 | 72 (54 - 86) | 0 (-1 — 1) | 1 |
| LPV: low priority | all | 170/33/52/85 | 72/22/32/46 | 1.2 (0.9 - 1.6) | 1 | 1.4 (0.9 - 2.1) | 1 | 1.0 (0.7 - 1.5) | 1 | 70 (56 - 84.9) | 2 (-5 - 2) | 1 |
| NPV: no priority | all | 612/101/199/312 | 267/75/135/158 | 1.1 (0.9 - 1.3) | 1 | 1.2 (0.9 - 1.4) | 1 | 1.1 (0.9 - 1.3) | 1 | 73 (55 - 86) | 1 (-1 — 1) | 1 |

### HPV, High-priority missense variants

The 107 HPVs were carried by 151 AD cases and 29 controls, and these associated with an overall 6.1-fold increased risk of AD (95%CI: 4.1–9.0, p=5.3x10^-24^). Specifically, HPVs associated with a 9.9-fold increased risk of EOAD (95%CI: 6.5–15.2, p=7.8x10^-29^), and to a 4.2-fold increased risk of LOAD (95%CI 2.7–6.5; p=1.3x10^-10^) (**Table 3**). Of the 107 HPVs, 77 (72%) were singletons (66/77 in AD cases), we observed 18 HPVs in two individuals (30/36 were AD cases), 12 variants in ≥3 individuals, and one variant (p.Y391C) in 12 individuals in the sample (all AD cases). The median AAO of HPV-carriers was 64 years (10%-90% range: 53-79), 8 years (95%CI -10– -6; 5.3x10^-9^) earlier than *SORL1* WT AD cases (**Figure 4A, Table S6**).

#### VPS10p-domain (res 82-617) and 10CC-domain (res 618-753)

The 27 HPVs in the VPS10p-domain associate with overall 8.8-fold increased risk of AD (95%CI 3.5–22.34; p=3.4x10^-7^); with a 15.7-fold increased risk of EOAD (95%CI 5.9–41.5; p=2.2x10^-9^) and a 5.5-fold increased risk of LOAD (95%CI: 2.0–15.0; p=9.5x10^-3^) (**Table 3**). We identified 6 HPVs involving cysteines in the L1 or L2 loops, which are involved in ligand-binding and unique to the VPS10p-domain of SORL1(36). Intriguingly, 12 unrelated AD cases (no controls) gained a cysteine in L1 (p.Y391C), suggesting that this variant is highly penetrant. Carriers had a median AAO of 67.5 years, which was 5.5 years later compared to PTV carriers, but 4.5 years earlier than *SORL1* WT carriers (**Fig 4B, Table S6**). One control (age <50) gained a cysteine in L1 (p.G398C), two cases lost, and two cases gained a cysteine in L2 (C467Y, C473S and twice R480C), and one control (age between 50-75) gained a cysteine in L1 (p.G398C, age <50) or L2 (p.S474C, age between 50-75). We further identified 16 HPVs with a REVEL score ≥0.50, carried by 16 cases and 3 controls, which in aggregate had an AAO of 2.5 years earlier than PTV-carriers (95%CI: -4–13) and 12.5 years earlier than *SORL1* WT carriers (**Fig 4B, Table S6**). Of particular interest are the four variants that affect the Asp-box, that stabilizes the β-propeller by forming interactions between propeller blades, with the L1/L2 loops and with the nearby 10CC-domains: three cases with p.S564G, p.T570I or p.S138F, and two cases with p.D236G. The 10CC-domain, C-terminal to the p.VPS10p-domain, stabilizes VPS10p β-propeller, and losing one of the 10 highly conserved cysteines or gaining a cysteine will likely impair domain folding. One case carried p.C716W, and two cases carried p.Y722C. In aggregate, we observed 19 variants affecting the 10CC-domain, with a median AAO of 67 years, 5 years earlier than *SORL1* WT carriers (**Fig 4B, Table S6**).

#### YWTD-domain (res 754-1013) and EGF-domain (res 1014-1074)

We identified 6 HPVs that affect the highly conserved YWTD-motif, which maintains the structural and functional integrity of the β-propeller. These were carried by 8 AD cases, with an AAO of 8 years earlier than *SORL1* WT carriers (**Fig 4B, Table S6)**. Five cases (AAO 46-78) and one control carried the deleterious p.R953H variant, substituting a positively charged arginine at position 38 in the 5^th^ YWTD-domain (see accompanying manuscript(35)). The EGF-domain includes eight cysteines that likely form four intradomain disulfide-bridges to stabilize the EGF:YWTD β-propeller unit (**Fig 1**). One case carried a p.Y1064C substitution and one control carried a p.C1026R substitution. Taken together, carrying a variant affecting the YWTD-domain leads to a 4.6-fold increased risk of AD (**Table 3)**.

#### CR-domain (res 1075-1550): calcium-cage and cysteines

13 cases and no controls carried 12 unique variants affecting calcium-cage residues, suggesting high penetrance. The median AAO of carriers was 12 years earlier than *SORL1 WT* cases, and 2 years *earlier* compared to PTVs carriers. (**Fig 4B, Table S6)**. One calcium-cage variant, p.D1108N, affecting the 1^st^ CR-domain, was observed in three unrelated AD cases (AAO 54-73). Another case (*APOE-ε3*/*ε3*, AAO <70) carried two calcium-cage variants, p.D1261G in the 5^th^ CR domain and p.D1345N in the 7^th^ CR domain; due to the distance between them on DNA level (∼3kb) we could not determine whether these variants were *in cis* or *in trans*. Furthermore, a disruption of the conserved pattern of 6 cysteines (odd number of cysteines, ONC) impairs CR-domain folding and leads to dysfunctional SORL1 protein. We identified 15 cysteine-gained and 7 cysteine-lost variants carried by 34 cases and 8 controls: such that carrying an ONC variant associates with a 4.9-fold increased risk of AD (95%CI: 2.3–10.6; p=1.8x10^-4^) (**Table 3**). Among the genetically unrelated individuals, we observed p.R1490C in five cases and two controls, p.R1080C in four cases, p.R1124C in three cases and one control. The median AAO of ONC-carriers was 68 years, 4 years earlier than *SORL1* WT-carriers, and 6-years *later* than PTV-carriers (**Fig 4B, Table S6**). One case (*APOE-ε3*/*ε4*, AAO <50) carried a p.C1453F ONC variant in combination with a PTV: variants were too far apart to confirm whether they were *in-cis* or *in-trans*.

#### 3FN-domain (res 1551-2121)

Disturbing domain-stabilizing interaction between the leucine at domain-position 77, a proline at position 79, and a tyrosine at position 83 (‘tyrosine corner’) critically impairs SORL1 dimerization(19). Six unrelated cases carried a p.Y1816C (position 83, average AAO 60.2 years), one case carried p.P1619Q (position 79, AAO <60). One <50 years old control carried p.L1617V (position 77): given the importance of the tyrosine corner in SORLA function, it is not unlikely that this individual will develop AD at a later age. Conserved glycines at positions 36 and 96 may affect domain stability or ligand binding: we identified one case with a p.G1732A (position 96) with AAO < 75 and one case (*APOE-*ε*2*/ε*4*) with a p.G1681D (position 36) with AAO <50. Lastly, four cases and one control carried variants affecting the residues that contribute to the hydrophobic core of the 3FN-domain, which acts as the ‘glue’ that connects the two β-sheets that constitute the 3Fn-domain sandwich (tryptophan at position 25 and the tyrosine at position 41). Furthermore, substitution of the moderate-priority prolines at positions 6, and 7 that occur in some 3FN-domains were observed only in cases.

#### Transmembrane and tail domain (res 2161-2214)

There were no variants prioritized in these domains.

### Effect of *APOE-ε4* allele

In the dataset (excluding the ADSP cohort, methods) AD risk increases 3.1-fold for each added *APOE*-*ε4* allele (95%CI 2.9–3.3, p=0). The median AAO for *APOE-ε4/ε4* AD cases was 64 years for those wild-type for *SORL1* (10%-90% range: 54-77), 60 years for PTV-carriers (10%-90% range: 53-65), and 58.5 years for HPV-carriers (10%-90% range: 53-69). The median AAO for *APOE-ε4* heterozygous AD cases was 70 years for those wild-type for *SORL1* (10%-90% range: 55-82), 61 years for PTV-carriers (10%-90% range: 51-74), and 63 years for HPV-carriers (10%-90% range: 54-78). The median AAO *APOE-ε4*-negative AD cases was 75 years for those wild-type for *SORL1* (10%-90% range: 56-89), 69 years for PTV-carriers (10%-90% range: 52-81), and 66 years for HPV carriers (10%-90% range: 48-86) (**Fig 5, Table S7**). Together, carrying a *SORL1* PTV or HPV expedited AAO by respectively 6 years (95%CI -10– -2) and 9 years (95%CI -13– -1.7) for *APOE-ε4*-negative AD cases, by respectively 9 (95%CI -10 – -6) and 7 (95%CI -8– -4) years for *APOE-ε4*-heterozyous cases, and by respectively 4 (95%CI -6–NA) and 5.5 (95%CI -8–NA) years for *APOE-ε4/ε4* cases. This indicates a major additive effect of APOE genotype. Evidence for an interactive effect in PTV- and HPV-carriers was limited (p=0.04 and p=0.06 respectively). However, inferences regarding a possible interaction effect may be incorrect as this case/control analysis design lacks power: only 18 controls with known *APOE* genotype carried a PTV or HPV, of whom 15 were negative for the ε4 allele (83%). Also, 5 were younger than 65 (28%) such that future AD is not unlikely in variant carriers. Nevertheless, the additive and possibly synergistic effect of *APOE-ε4* allele explains, in part, the variability in AAO of carriers of the same variant. The AAO-range of the twelve Y391C cases was 60–86 years, 56–91 years for five p.R744RX carriers, 60–73 years for four p.R866X carriers, 46–78 years for p.R953H carriers, and 56–74 for six p.Y1816C carriers (**Fig S3**). Indeed, EOAD cases were more likely to carry at least one *APOE*-*ε4* allele, while older cases often carried a protective *APOE*-*ε2* allele (**Fig S4**).

**Figure 5:**
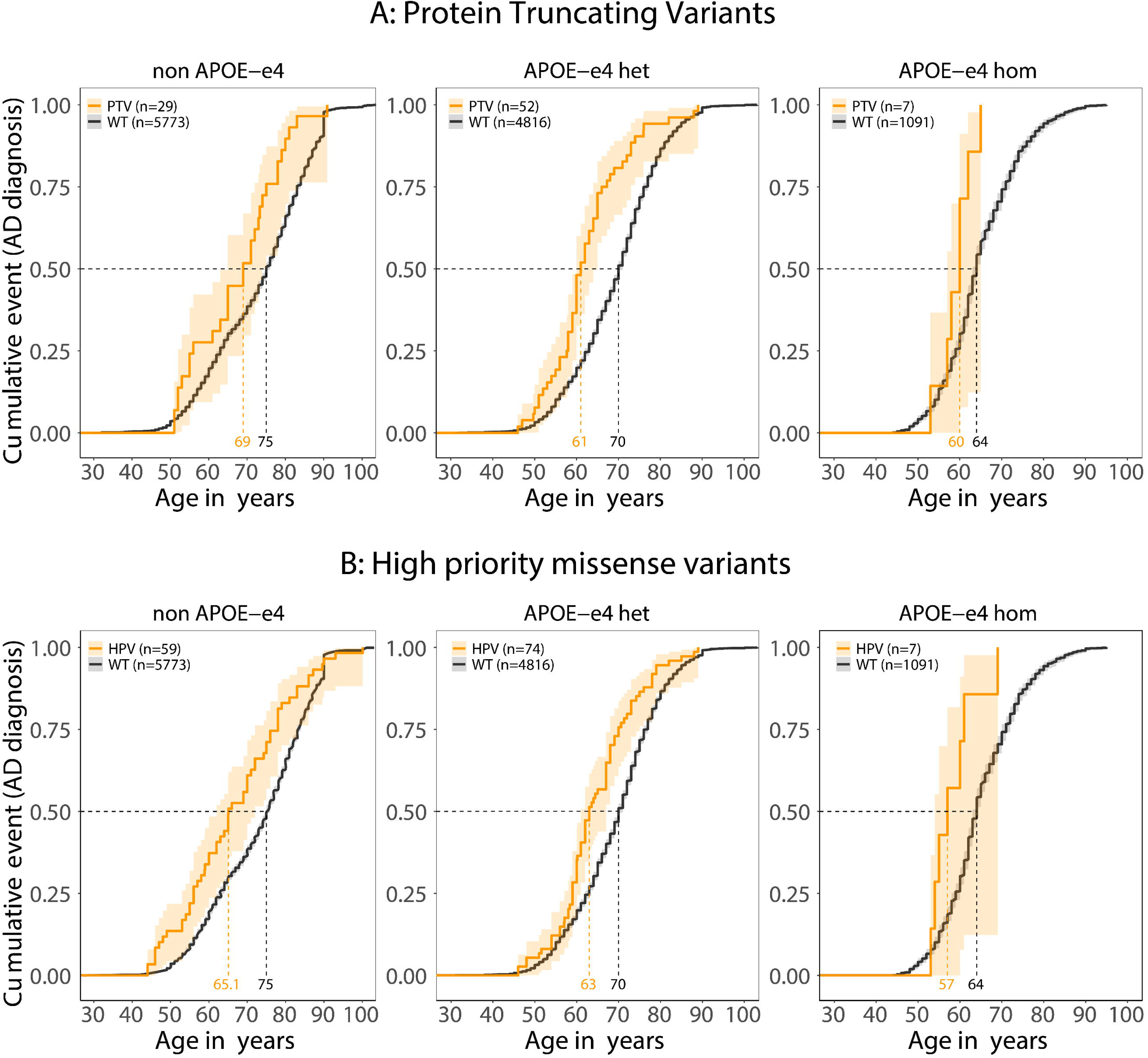
Rare *SORL1* variants in context of APOE genotype. **A.** AD cases who carried PTV in *SORL1* have an earlier age at onset compared to *SORL1* WT carriers with the same APOE genotype. The orange ages indicate at what age respectively 50% of the variant carriers had AD, the black ages indicate at the age at which 50% of the *SORL1* WT carriers developed AD. **B.** High-priority variants in context of APOE genotype. carriers of a high-priority variant have an earlier age at onset compared to wildtype carriers with similar APOE genotype. The effects of MPVs, LPVs and NPVs in context of APOE is shown in **Fig S2**.

### MPVs, LPVs, NPVs and variants with MAF>0.05%

MPVs, LPVs and NPVs had no or negligible effects on AD (**Fig 4A**, **Table 3**) and there was no change in AAO relative to *SORL1* WT cases (**Fig 4C)**. Likewise, the most common coding *SORL1* variants p.A528T and p.E270K with sample-MAFs of respectively 3.6% and 1.9%, are associated with a 1.11-fold and 1.02-fold increased risk of AD when imputed in GWAS(37). For p.D2065V, with sample-MAF 0.46%, we observed a 1.36-fold increased AD risk, and carriers had a slightly expedited AAO relative to *SORL1* WT cases (**Figure 4C**). See the **Supplement** for a more in-depth analyses of these variants.

### Comparison of prioritization scheme vs using only REVEL scores or AlphaMissense

While in aggregate, the 107 HPVs prioritized by DMDM(35), associated with a 6.1-fold increased risk of AD (i.e EAOD and LOAD combined), the aggregate of variants with a REVEL(32) or the AI-based AlphaMissense(38) score >0.9 (both with score range 0-1) was associated respectively with a 3.9-fold (95%CI 1.9-8.0) and 3.5-fold (95%CI 2.1-5.5) increased risk of AD (**Fig S5**). This indicates that manual variant annotation based on years of expertise, outperforms the utilization of *in silico* or AI-based prediction tools.

## DISCUSSION

In our assembled sample of 18,959 AD cases and 21,893 controls we identified 107 rare missense variants with MAF<0.05% as ‘high-priority’ HPV after applying a manual DMDM analysis(35). HPVs are associated with a 6-fold increased risk of overall AD (EOAD and LOAD combined), and 10-fold increased risk of EOAD. In this sample, the carriers of such variants had a median AAO of 64 years, 8 years earlier than carriers of wild-type *SORL1.* In comparison, carrying a PTV associated with an overall 17-fold increased risk of overall AD relative to non-carriers, and a 35-fold increased risk of EOAD. *SORL1* PTV-carriers in this sample had a median AAO of 62 years, 10 years earlier than carriers of wild-type *SORL1*. Other rare missense variants with MAF<0.05%, or coding variants with MAF>0.05% had no or only limited effects on AD risk.

Although the median AAO of *SORL1* PTV or HPV-carriers indicates a predisposition for EOAD, AAO is several years later than for carriers of established pathogenic variants in the *PSEN1*, *APP* , and *PSEN2* ADAD genes, with average AAO respectively ∼45, ∼50 years and ∼55 years(39–41). Akin to observations in established ADAD, we observe an additive (and possibly synergistic) effect of *APOE* genotype on AAO of *SORL1* PTV and HPV-carriers(42, 43). The median AAOs for PTV and HPV-carriers who are ε4/ε4, ε4-heterozygous or ε4-negative are respectively 60, 61 and 69 years and 58.5, 63 and 66 years, which is in agreement with a report stating that penetrance for *SORL1-*PTV and HPV-carriers combined was complete by age 70 among ε4/ε4 carriers, 10 years later for ε4-heterozygous carriers and even later for non-ε4 variant-carriers(14). In addition to the *APOE* genotype, other genetic risk factors will further influence AAO, akin to observations in established ADAD (37, 39, 43).

On average, HPVs and PTVs have a similar AAO. However, when grouping variants based on affected residues, certain HPVs have an earlier AAO than PTVs, while others have a later AAO. Variants that affect a calcium-cage residue in one of the CR-domains(44, 45), the YWTD-motif or the VPS10p-domain had an *earlier* AAO than carriers of PTV variants, suggesting they might have a dominant negative effect. Notably, variants affecting the YWTD-motif or a calcium-cage were observed only in AD cases, indicative of high penetrance. A dominant negative effect may be explained by requirement for SORL1 to dimerize (or possibly polymerize) at its 3FN-domains, before docking into the retromer(1, 46). Variants that lead to impaired protein folding cannot be properly matured at the endoplasmic reticulum (ER)(47), such that the receptor cannot exit the ER. However, so long as the 3FN-domains remain in-tact, any mutant SORL1 will retain the propensity to dimerize while still in the ER, not only with other mutant-SORL1, but also WT-SORL1. This way, *less* than half of SORL1 protein can exit the ER to perform its cargo trafficking functions. This mode of pathogenicity may also explain the (familial) AD observed for the carriers of the p.R953H and the p.R953C ‘Seattle variant’ affecting YWTD-domain folding(20), and the p.G511R variant, affecting VPS10p-domain folding(8). ER-retention prevents SORL1 protein to traffic to the cell surface, such that these variants will likely lead decreased shedding of soluble SORL1 (sSORL1) in the interstitial space(48). We acknowledge that the earlier AOO of these specific *SORL1* missense variants relative to PTVs concerns only few carriers, such that significance cannot be reached. Therefore, the associated dominant negative effects need to be confirmed in an independent large sequencing dataset of AD cases and controls and/or by further functional studies.

The effect on AAO of other HPVs may be similar to the haploinsufficiency associated with PTVs. An example is the p.Y1816C mutant that affects the ‘tyrosine corner’, a residue that contributes strongly to the stability of 3^rd^ 3FN-domain(19) which was carried by the probands and affected family members of three unrelated pedigrees(19). Functional experiments indicated that the p.Y1816C mutant is efficiently matured and trafficked from the ER to the endosome. Once there, it fails to form the dimer-dependent complex with retromer, such that the SORL1 mutant cannot contribute to retromer sorting. However, the wild type allele still can, suggesting that the variant leads to haploinsufficiency. We observed that sSORL1 levels were decreased to ∼50% for carriers of this variant, mimicking the effect we observe for PTVs on sSORLA shedding(49). This mode of pathogenicity may also apply to the p.G1732A variant, likely impairing the folding of the 2^nd^ 3FN-domain, which was identified in a pedigree affected with EOAD(17).

Other HPVs lead to a slightly *later* AAO than PTV variant carriers, suggesting that their effects are less damaging than losing one *SORL1* copy. We speculate that the p.Y391C substitution affecting Loop L1 in the VPS10p-domain (observed in 12 cases) affects ligand-binding of the VPS10p-domain, leading to decreased lysosomal delivery of (among other ligands) Amyloid-β and to an increase of secreted Amyloid-β(50). However, other SORL1 functions may still be in-tact, which may explain a less deleterious effect compared to carrying a PTV. Carriers of variants affecting the 10CC-domain, or that lead to losing or gaining a cysteine in the CR-domain also have a later AAO compared to PTV carriers, suggesting some residual activity for these mutants. These variants may lead to an unstable receptor, some of which may be removed by the ER-associated degradation pathway, while others may escape the ER control-check and be exported to subsequent cellular compartments. While we cannot provide any supporting evidence at current, this mode of pathogenicity might explain the EOAD observed for carriers of the p.R1303C substitution (cysteine-loss in the 6^th^ CR-domain), in one of the pedigrees described by Thonberg *et al.*(17). Additional functional evidence is necessary to support the different modes of pathogenicity associated with different HPV-types.

Ideally, risk and AAO analysis are assessed in population-based follow-up studies. However, genetic variants associated with EOAD are extremely rare, such that sequencing all individuals from a population study will not yield informative data. Therefore, our dataset is an assembled sample of AD cases and controls which is relatively enriched with EOAD cases. The effect of this enrichment is observable in the earlier median AAO of AD cases that are homozygous, heterozygous or negative for the *APOE-ε4* allele (respectively 64, 70 and 75 years) compared to a population sample (70, 74.5, 82 years, as estimated from Reiman *et al*.(51)). We acknowledge that determining odds ratios for EOAD and LOAD separately only partly accounts for the influence of age on the effect of *SORL1* variants on AD risk. Note also, that all effect sizes were calculated relative to the same control group and that at the time of sample inclusion, 50% of the controls was younger than 65 (33% was even younger than 50), making it likely that some controls may develop AD at a later age, such that effect sizes presented here may be conservative. On the other hand, recent analyses have indicated that within a pedigree, the AAO of the index patient was significantly earlier than those of family-members(14). We suspect that our sample of genetically unrelated individuals may be enriched with index patients, which may skew the distribution towards relatively early AAO. Nevertheless, it is valid to compare effects on AD risk and AAO distributions between the different *SORL1* variants *within* our sample, as similar biases apply to all other AD cases and controls. Overall, we caution that AAO distributions and effects on AD risk are not representative of the overall population.

Several *SORL1*-features may be considered when investigating *SORL1* variants. Exons 23-33 each translate one CR-domain, such that exon-skipping splice-variants may translate to a SORL1 protein lacking one CR-domain. It is unclear whether this yields an inactive or (partly) active SORL1(52). An exception is the 7^th^ CR-domain, encoded by exon 29, since joining exons 28 and 30 produces a nonsense-codon. Exon 1 was excluded from analysis due to differential missingness, however upon inspection we identified 9 PTVs in exon 1, of which 4 in controls, while PTVs in the rest of the gene occurred almost exclusively in AD cases. Although we cannot provide supporting evidence to substantiate this, the use of an alternative transcriptional start site or Nonsense-mediated decay escape may be a back-up mechanism for *SORL1* transcription(53). Furthermore, it is unclear whether variants observed in several genetically unrelated individuals share a founder mutation: p.Y391C (12 cases), p.R1490C (5 cases/2 controls), p.Y1816C (6 cases), p.R953H (6 cases/1 control), p.R744X (5 cases) and p.R866X (4 cases). If these variants occurred *de novo* in each pedigree this might provide preliminary evidence for mutation hotspots in *SORL1*.

## Conclusion

Here, we show that carriers of *SORL1* PTVs and HPVs have diverse AAOs, which are influenced by *APOE* genotype, complicating penetrance estimates(14, 54). This is in parallel with variants in genes implicated in hereditary breast cancer including the ‘high risk’ genes *BRCA1*, *BRCA2,* and *PALB2* and ‘moderate risk’ genes *CHEK2* and *ATM*(*55*). In all these genes, PTVs generally associate strongest with disease risk while missense variants have variable effects and are notoriously more difficult to classify. The oncogenetics field has created specific guidelines for testing and counseling high- and moderate risk genes,(56) which may be explained by the actionability of oncogenes (e.g., screening programs or preventive surgery). In the absence of actionability, the AD field has historically been more hesitant to adopt genetic testing of genes for which variants that segregate with AD across multiple generations have not yet been identified. Nevertheless, some historically high-risk genes for neurodegenerative disorders, such as variants in *NOTCH3* that cause CADASIL (Cerebral Autosomal Dominant Arteriopathy with Subcortical Infarcts and Leukoencephalopathy), are now considered a spectrum of high and moderate risk conferring variants, and they are counseled accordingly. Here, we show that a subset of *SORL1* variants is highly penetrant for AD with an AAO that overlaps with that observed for carriers of known pathogenic variants in *PSEN2*(*23*) and even some variants in *PSEN1*(57). With actionability of Alzheimer’s Disease on the horizon, our results encourage engaging a discussion on whether reporting these variants to patient-carriers is desirable, possibly in combination with *APOE* genotype.

## List of abbreviations

AAO: Age at onset
AD: Alzheimer’s disease
ADAD: Autosomal dominant Alzheimer’s disease
APP: Amyloid precursor protein
DMDM: Domain-mapping of disease mutations
EOAD: Early onset Alzheimer’s disease
ER: Endoplasmic Reticulum
GWAS: Genome wide association Studies
HPV: High priority variant
LOAD: Late onset Alzheimer’s disease
LPV: Low priority variant
MAF: Minor allele frequency
MPV: Moderate priority variant
NPV: No priority variant
ONC: Odd number Cysteines
PCA: Principal Component Analysis
PTV: Protein truncating variants
REVEL: rare exome variant ensemble learner
VEP: variant effect predictor
VUS: variant of unknown significance
WES: Whole exome sequencing
WGS: Whole genome sequencing
WT: wild-type

## Declarations

### Ethics approval and consent to participate

For consent and sample description see supplementary data 1.1.

### Consent for publication

Not applicable.

### Availability of data and materials

All 646 identified *SORL1* coding variants are listed in the recent release of the Alzforum Mutation database(58). (https://www.alzforum.org/mutations/sorl1). Genomes from contributing cohorts are shared on the Alzheimer Genetics Hub: www.alzheimergenetics.org.

### Author contributions

Conceived the study: HH^1^ and OMA; Wrote the manuscript: HH^1^, MdW, NT, SvdL, MH, OMA; Genetic Analyses: HH^1^, MdW, NT, SvdL, CdG, MV, RvS, MH, OMA Participant collection and sequencing from the ADES, ADSP, StEP-AD, Knight ADRC, UCSF/NYGC/UAB cohorts: SA, NA, PA, GWB, CB^1^, CB^2^, JCB, AB, PB, FB, JB, DC, CC^1^, JC, JNC, CC^2^; AD, J-FD^1^, SD, J-FD^2^; ND, ALDS, OD-I, CMvD, LAF, MVF, WMvdF, NCF, DG^1^, EG; JJPG, MG, BG-B, DG^2^, YLG, RG, JLH, CH, HH^2^; MAI; MKI; IEJ, AK, RK, J-CL, ML; AWL, AL, LL, MAMM, RM, ERM, CM, RM, SM, PM, AM, MOM, KM, RMM, BN, ACN, VN, GN, PJN, FP, PP, MAP-V, YALP, OQ, AR, RR^1^; RR^2^, MJTR, A-CR, SGR-H, FR, SR, JGJvR, NSR, SS^1^, PS-J, GDS, PS, JMS, DS^1^, SS^2^, DS^2^; RS, EAS, SS^3^, GS, JCvS, BT, AGU, PJV, MW, DW, L-SW, JW, JSY, AZ. CB^1^: Céline Bellenguez; CB2: Claudine Berr; CC^1^: Camille Charbonnier; CC^2^: Carlos Cruchaga; J-FD^1^: Jean-François Dartigue; J-FD^2^: Jean-François Deleuze; DG^1^: Daniela Galimberti; DG^2^: Detelina Grozeva; HH^1^ Henne Holstege; HH^2^ Holger Hummerich; RR^1^: Rachel Raybould; RR^2^: Richard Redon; SS^1^: Salha Saad; SS^2^: Sudha Seshadri; SS^3^: Sandro Sorbi

## Data Availability

All data produced in the present work are contained in the manuscript.

## Acknowledgments

The authors are grateful to all study participants, their family members, the participating medical staff, general practitioners, pharmacists and all laboratory personnel involved in patient diagnosis, blood collection, blood biobanking, DNA preparation and sequencing. We further acknowledge the collaborative support of the Alzforum, specifically Elizabeth Wu and Kathleen Zahs in this work, for critical feedback of the prioritization approach and for making all variants available on the Alzforum SORL1 database (https://www.alzforum.org/mutations/sorl1). The data used in this work was collected using the funding obtained by the following study cohorts: ADES-FR, AgeCoDe-UKBonn; Barcelona SPIN; AC-EMC; ERF and Rotterdam; ADC-Amsterdam; 100-plus study; EMIF-90+; Control Brain Consortium; PERADES; StEP-AD; Knight-ADRC; UCSF/NYGC/UAB; UCL-DRC EOAD; ADSP. Data used in preparation of this article were obtained from the Alzheimer’s Disease Neuroimaging Initiative (ADNI) database (https://adni.loni.usc.edu/). Full consortium acknowledgements and funding sources are listed in the Supplementary Note.

The work in this manuscript was carried out on the Cartesius supercomputer, which is embedded in the Dutch national e-infrastructure with the support of SURF Cooperative. Computing hours were granted to H. H. by the Dutch Research Council (‘100plus’: project# vuh15226, 15318, 17232, and 2020.030; ‘Role of VNTRs in AD’; project# 2022.31, ‘Alzheimer’s Genetics Hub’ project# 2022.38).

