## Supplemental Data for "Domain mapping of disease mutations supports genetic testing of specific *SORL1* variants in familial Alzheimer’s Disease"

TO:

1. Department of Human Genetics, Amsterdam UMC, location Vrije Universiteit Amsterdam, Amsterdam, The Netherlands
2. Alzheimer Center Amsterdam, Neurology, Amsterdam UMC, location Vrije Universiteit Amsterdam, Amsterdam, The Netherlands
3. Amsterdam Neuroscience, Neurodegeneration, Amsterdam, The Netherlands
4. Delft Bioinformatics Lab, Delft University of Technology, Delft, The Netherlands
5. Clinical Genetics, Human Genetics, Amsterdam UMC, Amsterdam, The Netherlands
6. Genome Analysis Laboratory, Human Genetics, Amsterdam UMC, location Vrije Universiteit Amsterdam, the Netherlands
7. Department of Epidemiology, Erasmus Medical Centre, Rotterdam, The Netherlands
8. LACDR, Leiden, The Netherlands
9. Nuffield Department of Population Health Oxford University
10. Univ. Lille, Inserm, CHU Lille, Institut Pasteur Lille, LabEx DISTALZ-U1167-RID-AGE - Facteurs de risque et déterminants moléculaires des maladies liées au vieillissement, Lille, France
11. The John P. Hussman Institute for Human Genomics, University of Miami, Miami, Florida, USA
12. Univ Montpellier, Inserm, INM (Institute for Neurosciences of Montpellier), Montpellier, France
13. Cardiovascular Health Research Unit, Department of Medicine, University of Washington, Seattle, WA, USA
14. Université Paris-Saclay, CEA, Centre National de Recherche en Génomique Humaine Evry, France
15. Experimental Neuro-psychobiology Laboratory, Department of Clinical and Behavioral Neurology, IRCCS Santa Lucia Foundation, Rome, Italy
16. Department of Neurodegenerative Science, Van Andel Institute, Grand Rapids, MI, USA
17. Division of Psychiatry and Behavioral Medicine, Michigan State University College of Human Medicine, Grand Rapids, MI, USA
18. Univ Rouen Normandie, Normandie Univ, Inserm U1245 and CHU Rouen, Departments of Genetics and CNRMAJ, F-76000 Rouen, France
19. Memory Unit, Neurology Department and Institut de Recerca Sant Pau, Hospital de la Santa Creu i Sant Pau, Universitat Autònoma de Barcelona, Sant Quintí 77-79, 08041, Barcelona, Spain
20. CIBERNED, Network Center for Biomedical Research in Neurodegenerative Diseases, National Institute of Health Carlos III, Madrid, Spain
21. Neurogenomics and Informatics Center, Washington University School of Medicine, St Louis, MO, USA
22. Psychiatry Department, Washington University School of Medicine, St Louis, MO, USA
23. Hope Center for Neurological Disorders, Washington University School of Medicine, St Louis, MO, USA
24. Department of Neuroscience, Catholic University of Sacred Heart, Fondazione Policlinico Universitario A. Gemelli IRCCS, Rome, Italy
25. University Bordeaux, Inserm, Bordeaux Population Health Research Center, France
26. Department of Neurology, Bordeaux University Hospital, Bordeaux, France
27. UKDRI at Cardiff, School of Medicine, Cardiff University, Cardiff, UK
28. Department of Biostatistics, Boston University School of Public Health, Boston, MA, USA
29. Framingham Heart Study, Framingham, MA, USA
30. Department of Neurology, Boston University School of Medicine, Boston, MA, USA
31. Department of Epidemiology, Boston University, Boston, MA, USA
32. Department of Medicine (Biomedical Genetics), Boston University, Boston, MA, USA
33. Dementia Research Centre, UCL Queen Square Institute of Neurology, London, UK
34. UK Dementia Research Institute at UCL, London, UK
35. Fondazione IRCCS Ca' Granda, Ospedale Policlinico, Milan, Italy
36. University of Milan, Milan, Italy
37. Univ Brest, Inserm, EFS, CHU Brest, UMR 1078, GGB, F-29200, Brest, France
38. Division of Psychological Medicine and Clinical Neuroscience, School of Medicine, Cardiff University, Cardiff, UK
39. Quantitative Sciences Unit, Department of Medicine, Stanford University, Stanford, CA, USA
40. Department of Population and Quantitative Health Sciences, School of Medicine, Case Western Reserve University, Cleveland, Ohio, USA
41. Clinical and Experimental Science, Faculty of Medicine, University of Southampton, Southampton, UK
42. MRC Prion Unit at UCL, UCL Institute of Prion Diseases, London, UK
43. Division of Neurogenetics and Molecular Psychiatry, Department of Psychiatry and Psychotherapy, Faculty of Medicine and University Hospital Cologne, University of Cologne, Cologne Germany
44. Department of Internal Medicine, Erasmus Medical Centre, Rotterdam, The Netherlands
45. McGill University and Genome Quebec Innovation Centre, Montreal, QC, Canada
46. HudsonAlpha Institute for Biotechnology, Huntsville, AL, USA
47. Department of Human Genetics, Amsterdam UMC, University of Amsterdam, Amsterdam Reproduction and Development Research Institute Amsterdam, The Netherlands
48. Dr. John T. Macdonald Foundation Department of Human Genetics, University of Miami, Miami, Florida, USA
49. Institute of Neurology, Catholic University of the Sacred Heart, Rome, Italy
50. Taub Institute on Alzheimer's Disease and the Aging Brain, Department of Neurology, Columbia University, New York, New York, USA
51. Gertrude H. Sergievsky Center, Columbia University, New York, NY, USA
52. Division of Gerontology and Geriatrics, Department of Medicine and Surgery, University of Perugia, Perugia, Italy
53. Division of Clinical Geriatrics, Department of Neurobiology, Care Sciences and Society, Karolinska Institutet, Stockholm, Sweden
54. Department of Clinical Genetics, Erasmus Medical Center, Rotterdam, The Netherlands
55. Department of Neuroscience, Psychology, Drug Research and Child Health University of Florence, Florence, Italy
56. IRCCS Fondazione Don Carlo Gnocchi, Florence, Italy
57. Penn Neurodegeneration Genomics Center, Department of Biostatistics, Epidemiology, and Informatics; University of Pennsylvania Perelman School of Medicine, Philadelphia, PA, USA
58. Penn Neurodegeneration Genomics Center, Department of Pathology and Laboratory Medicine, University of Pennsylvania Perelman School of Medicine, Philadelphia, PA, USA
59. Genomic And Molecular Epidemiology (GAME) Lab, School of Biosciences and Veterinary Medicine, University of Camerino (UNICAM) Camerino, 62032, Italy
60. Univ. Lille, Inserm, CHU Lille, UMR1172, Resources and Research Memory Center (MRRC) of Distalz, Licend, Lille France
61. Unit of Neurodegenerative diseases, Department of Neurology, University Hospital Germans Trias i Pujol, Badalona, Barcelona, Spain
62. The Germans Trias i Pujol Research Institute (IGTP), Badalona, Barcelona, Spain
63. Laboratory of Neuropsychiatry, Department of Clinical and Behavioral Neurology, IRCCS Santa Lucia Foundation, Rome, Italy
64. Department of Psychiatry and Glenn Biggs Institute for Alzheimer’s and Neurodegenerative Diseases, San Antonio, TX, USA
65. Department of Neurodegenerative Diseases and Geriatric Psychiatry, University Hospital Bonn, Medical Faculty, Bonn, Germany
66. German Center for Neurodegenerative Diseases (DZNE, Bonn), Bonn, Germany
67. Cluster of Excellence Cellular Stress Responses in Aging-Associated Diseases (CECAD), University of Cologne, Cologne, Germany
68. Université de Nantes, CHU Nantes, CNRS, INSERM, l'institut du thorax, Nantes, France
69. Institute of Social Medicine, Occupational Health and Public Health, University of Leipzig, Leipzig, Germany
70. Neurology Service, Marqués de Valdecilla University Hospital (University of Cantabria and IDIVAL), Santander, Spain
71. Amsterdam Reproduction & Development research institute, Amsterdam UMC, Amsterdam, The Netherlands
72. Univ Rouen Normandie, Normandie Univ, Inserm U1245 and CHU Rouen, Departments of Neurology and CNRMAJ, F-76000 Rouen, France
73. MRC UK Dementia Research Institute, Division of Psychological Medicine, Cardiff University, Cardiff, UK.
74. Memory and Aging Center, Department of Neurology Weill Institute for Neurosciences, and Department of Radiology and Biomedical Imaging, University of California, San Francisco, CA, USA.
75. Dept. of Biomedicine, Aarhus University, Denmark

**INDEX**

[2.2.1	Table S1. XLS FILE: Rare variants (MAF < 0.05%)	26](#_Toc163552706)

### 1 Supplementary Data

#### 1.1 Sample Description

We analyzed a total sample of 52,361 individuals (40,852 after QC) sequenced with Illumina technology, collected as part of the Alzheimer Disease European Sequencing consortium (ADES), comprising 15 studies from Germany, France, The Netherlands, Spain, Italy, the United Kingdom and the USA (majority of which were from the Alzheimer’s Disease Sequencing Project (ADSP)^1^. All studies were approved by the ethics committees of respective institutes, and all participants provided informed consent for study participation.

Across all studies, AD cases were defined according to NIAA criteria^2^ for possible or probable AD or according to NINCDS-ADRDA criteria^3^ depending on the date of diagnosis. When possible, supportive evidence for an AD pathophysiological process was sought (including CSF biomarkers) or the diagnosis was confirmed by neuropathological examination (**Table**). Cases were annotated with the age at onset or age at diagnosis otherwise, samples were classified as late onset AD. Controls were not diagnosed with AD.

##### 1.1.1 ADES-FR

The ADES-FR project combines WES and WGS data from AD cases and controls from France^4^. Part of the patients are from the CNRMAJ-Rouen center (n=921) and patient ascertainment is described in detail in Nicolas et al.^5^ including an update of the inclusions by the French National network CNR-MAJ (national reference center for young Alzheimer patients). Briefly, unrelated cases with early-onset AD (age at onset ≤65 years) from France were recruited among patients who fulfilled the NIAA criteria^2^. The clinical examination included personal medical and family history assessment, neurologic examination, neuropsychological assessment, and neuroimaging. In addition, cerebrospinal fluid (CSF) biomarkers indicative of AD were available for 67% of the cases. Cases with CSF biomarkers not consistent with AD diagnostics were excluded. A positive family history (i.e., at least a secondary case among first- or second-degree relatives, whatever the age of onset) was present in 45% of cases. Patients were either screened by Sanger sequencing and QMPSF for pathogenic variants in *APP*, *PSEN1* or *PSEN2* prior to WES or by the interpretation of WES data or both. Carriers of pathogenic variants were not included for WES or were secondarily excluded following WES analysis so that none of the CNRMAJ-Rouen patients included in this work prior to shared analyses is a carrier of a pathogenic variant in *APP*, *PSEN1*, *PSEN2* as well as in a list of Mendelian dementia causative genes^6^. In addition, some controls were recruited directly from the CNRMAJ (n=30). Another large part of the samples was from the European Alzheimer’s Disease Initiative (EADI) dataset^7^. This study combined clinical prevalent and incident cases of AD (n=1,121) (i) from Lille cross-sectional studies and (ii) from the Three-City (3C) study, a population-based, prospective study with 12-years of follow-up^8^. Diagnoses were established according to the DSM-III-R and NINCDS-ADRDA criteria^3^. Controls were selected among the 3C individuals not diagnosed with dementia after a 12-year follow-up (n=670). In addition, other controls were obtained from the FREX consortium^9^. These controls (n=576) were specifically designed from 6 French cities with the aim of studying and establishing the French population genetic structure of rare variants. Overall, the ADES-FR samples includes 2,042 AD cases (1,088 EOAD and 954 LOAD) and 1,276 controls. All patients and controls provided informed written consent for genetic analyses in a clinical and/or in a research setting, according to each study. In addition, the ethics committee of the Rouen University Hospital approved the use of retrospective data in the context of the ADES-FR project and with other ADES European and American partners (CERNI notifications 2017-015 and 2019-055).

Furthermore, entire exomes of 529 independent and unrelated AD patients, including 384 patients from the ECASCAD study were included. All had CSF biomarkers consistent with AD (except two patients who had neuropathological confirmation), and 90% of them were EOAD cases, the remaining 10% cases had an age of onset between 65 and 75 years. As controls, we used the extracted BAM files of the SORL1 gene among the genome sequencing data from the FranceGenRef study. Individuals included in this study were selected based on the places of birth of their grandparents within France and at a maximum distance of 30 kilometers. A total of 862 individuals (274 females and 588 males) were sampled from three different studies: 50 individuals (25 females and 25 males) were blood donors sampled in the Finistère district, 354 individuals (177 females and 177 males) were blood donors from the PREGO biobank with ancestries in the other districts of Brittany (Côtes d’Armor, Ile-et-Vilaine, Morbihan) and in the 5 districts of the Pays-de-la-Loire region (Loire-Atlantique, Maine-et-Loire, Mayenne, Sarthe, Vendée), 458 individuals (72 females and 386 males) were volunteers from the GAZEL cohort (www.gazel.inserm.fr/en) who were selected among the volunteers who gave a blood sample and who answered a questionnaire on their parents and grandparents’ places of birth. All individuals signed informed consent for genetic studies at the time they were enrolled and had their blood collected.

##### 1.1.2 AgeCoDe-UKBonn

The AgeCoDe-UKBonn sample was derived from the following two sources, the German study on Aging, Cognition, and Dementia in primary care patients (AgeCoDe, n=294) and the interdisciplinary Memory Clinic at the University Hospital of Bonn (UKBonn, n=100).

—**The German study on Aging, Cognition, and Dementia**: The AgeCoDe study is a multicenter prospective general practice-based cohort study since 2001, including community dwelling elderly aged 75 years or older that were recruited at six study sites (Bonn, Düsseldorf, Hamburg, Leipzig, Mannheim, and Munich). The AgeCoDe study was approved by the local ethics committees of the Universities of Bonn, Hamburg, Düsseldorf, Heidelberg/Mannheim, Leipzig, and Munich. Before participation written informed consents were collected from all subjects. The AgeCoDe study aims to identify risk factors and predictors of cognitive decline and dementia^10,11^. Participants were recruited from general practitioner (GP) registries. Inclusion criteria were an age of 75 and older, absence of dementia, one or more visits to the GP in the past year, no hearing or vision impairments and German as a native language. Exclusion criteria were only home-based GP consultations, severe illness with a fatal outcome within 3 months and a language barrier. The baseline assessment including 3,327 subjects was completed between 2002 and 2003. After the baseline assessment 70 subjects were excluded due to presence of dementia after standard assessment and 40 subjects were excluded with an age below 75 years. Participants were interviewed for follow up every 18 months. All assessments are performed at the participant’s home by a trained study psychologist or physician. At all visits, assessment includes the Structured Interview for Diagnosis of Dementia of Alzheimer type, Multi-infarct Dementia, and Dementia of other etiology according to DSM-IV and ICD-10 (SIDAM)^12^. The SIDAM comprises: (1) a 55-item neuropsychological test battery, including all 30 items of the MMSE and assessment of several cognitive domains (orientation, verbal and visual memory, intellectual abilities, verbal abilities/ calculation, visual–spatial constructional abilities, aphasia/ apraxia); (2) a 14-item scale for the assessment of the activities of daily living (SIDAM-ADL-Scale); and (3) the Hachinski Rosen-Scale. Dementia was diagnosed according to DSM-IV criteria. AgeCoDe provided DNA from 294 persons who progressed to late onset AD dementia at any follow up.

—**UKBonn**: The interdisciplinary Memory Clinic of the Department of Psychiatry and Department of Neurology at the University Hospital in Bonn provided early-onset AD patients (n=100). Diagnoses were assigned according the NINCDS/ADRDA criteria^3^ and on the basis of clinical history, physical examination, neuropsychological testing (using the CERAD neuropsychological battery, including the MMSE), laboratory assessments, and brain imaging.

##### 1.1.3 Barcelona- SPIN

Neuropathological samples were obtained from the Neurological Tissue Bank of the Biobanc-HospitalClinic-IDIBAPS, and disease evaluation was performed according to international consensus criteria. Clinical samples were recruited from the multimodal Sant Pau Initiative on Neurodegeneration (SPIN) cohort (<https://santpaumemoryunit.com/our-research/spin-cohort/>)^13^, and were evaluated at the Memory Unit at Hospital de Sant Pau (Barcelona). The repository includes clinical data of more than 6,000 participants, >2900 plasma samples, genetic material (DNA and RNA) of >3,200 and >400 subjects, respectively, and >2,000 CSF samples. All controls had normal cognitive scores in the formal neuropsychological evaluation and normal core CSF AD biomarkers, based on previously published cut-offs^14^. AD patients fulfilled clinical criteria of “probable AD dementia with evidence of the AD pathophysiological process”^3^ and therefore had abnormal core AD biomarkers (low Aβ1–42 and high t-Tau or p-Tau) in the CSF. The original protocol and the subsequent amendments were approved by our local Ethics Committee at the Sant Pau Research Institute as well as the Committee of the Neurological Tissue Bank. The SPIN cohort is based on blinded enrollment and only clinically relevant biomarker results are disclosed.

##### 1.1.4 AC-EMC

The Alzheimer Center Erasmus MC cohort (AC-EMC) includes patient referred to the Department of Neurology of the Erasmus Medical Center (Rotterdam, the Netherlands). DNA samples from 125 patients with probable AD were included in the current study. The average age at onset was 60 years (range 41-77). A large fraction of the patients had a positive family history, defined as at least one first degree relative with dementia. All patients underwent clinical examination, neuropsychological assessment, neuroimaging, and if indicated, a lumbar puncture. The diagnosis was established according to the National Institute of Neurological and Communicative Disorders and Stroke-Alzheimer’s Disease and Related Disorders Association (NINCDS-ADRDA) criteria for AD^3^.The study was approved by the Medical Ethical Committee of the Erasmus Medical Center, and written informed consent was obtained from all participants or their legal representatives.

##### 1.1.5 ERF

The Erasmus Rucphen Family (ERF) Study is a family-based cohort study that is embedded in the Genetic Research in Isolated Populations (GRIP) program in the South West of the Netherlands. The aim of this program was to identify genetic risk factors in the development of complex disorders. For the ERF study, 22 families that had at least five children baptized in the community church between 1850-1900 were identified with the help of genealogical records. All living descendants of these couples and their spouses were invited to take part in the study. Data collection started in June 2002 and was finished in February 2005.

##### 1.1.6 Rotterdam Study

The Rotterdam Study^15^ is an ongoing prospective population-based cohort study, focused on chronic disabling conditions of the elderly^16^ of which a random subset was exome sequenced. Participants were screened for dementia at baseline and at follow-up examinations using the Mini-Mental State Examination (MMSE) and the Geriatric Mental Schedule (GMS) organic level^17^. Screen-positives (MMSE <26 or GMS organic level >0) underwent extensive examination^18^. Finally, individuals were diagnosed in accordance with standard criteria for dementia (Diagnostic and Statistical Manual of Mental Disorders, Third Edition, Revised (DSM-III-R)) and Alzheimer’s disease, NINCDS-ADRDA^3^. Follow-up for incident dementia was complete until January 1st, 2014. The Rotterdam Study has been approved by the Medical Ethics Committee of the Erasmus MC and by the Ministry of Health, Welfare and Sport of the Netherlands, implementing the Wet Bevolkingsonderzoek: ERGO (Population Studies Act: Rotterdam Study). All participants provided written informed consent to participate in the study and to obtain information from their treating physicians.

##### 1.1.7 ADC-Amsterdam

The ADC-Amsterdam cohort includes patients who visit the memory clinic of the Alzheimer Center at the Amsterdam University Medical Center, The Netherlands, and was described previously^19^. DNA samples from 854 patients with probable and possible AD were included in the current study. Additionally, 353 individuals diagnosed with psychiatric and subjective cognitive complaints were included as controls. Individuals in this cohort were extensively characterized to reduce the chance of misdiagnosis. Patients underwent an extensive standardized dementia assessment, including medical history, informant-based history, a physical examination, routine blood and CSF laboratory tests, neuropsychological testing, electroencephalogram (EEG) and MRI of the brain. The diagnosis of probable AD was based on the clinical criteria formulated by the National Institute of Neurological and Communicative Disorders and Stroke—Alzheimer’s Disease and Related Disorders Association (NINCDS-ADRDA) and based on National Institute of Aging–Alzheimer association (NIA-AA)^2^. Clinical diagnosis is made in consensus-based, multidisciplinary meetings. All patients gave informed consent for biobanking and for the use of their clinical data for research purposes. Selection for whole exome sequencing was based on an early age-of-onset (age at diagnosis <70 years) and available CSF biomarkers.

##### 1.1.8 Netherlands Brain Bank

From the Netherlands Brain Bank^20^ we selected brain tissues donated by patients diagnosed with Alzheimer Disease. DNA was isolated and used for WES sequencing.

##### 1.1.9 Amsterdam-UMC

This cohort consists of WES data that were generated as part of a diagnostic work-up. All samples are from healthy adults for whom WES analysis was performed to aid the analysis of a patient, in most cases these were healthy parents of an affected child for whom trio-WES analysis was performed. These parents either have no pathogenic variant, or are carrier of one recessive pathogenic variant that does not affect health.

##### 1.1.10 100-plus Study

The 100-plus Study, is a prospective cohort study of cognitively healthy centenarians that associated with the Alzheimer Center at the Amsterdam University Medical Center. Detailed participant recruitment and procedures were described previously^21^. Trained researchers visited the centenarians at their home residence annually, where they were subjected to questionnaires regarding demographics, lifestyle, medical history, physical well-being and objective measurements of cognitive and physical functions. Cognitive function is tested by an extensive neuropsychological testing battery. For the current study, DNA samples 375 centenarians were included who completed at least one neuropsychological test at baseline, and exome sequencing from 349 centenarians passed QC (removal was mostly due to kinship). Centenarians who scored >22 on the MMSE were regarded as controls, while centenarians who scored ≤22 were regarded as cases^22^. The Medical Ethics Committee of the Amsterdam UMC approved this study and informed consent was obtained from all participants. The study has been conducted in accordance with the declaration of Helsinki.

##### 1.1.11 EMIF-AD 90-plus Study

The EMIF-AD 90+ study^23^ is a cohort-study of the oldest-old (90+), situated at the Amsterdam UMC and the University of Manchester. The study contributed n=72 controls. Controls were tested to have a Mini-Mental State Examination (MMSE) >=26 and a global Clinical Dementia Rating (CDR) score of 0 at baseline.

##### 1.1.12 CBC: Control Brain Consortium

The Control Brain Consortium was previously described^24^. It consists of whole-exome sequencing in 478 samples derived from several brain banks in the United Kingdom and the United States of America. Samples were included when subjects were, at death, over 60 years of age, had no signs of neurological disease and were subjected to a neuropathological examination, which revealed no evidence of neurodegeneration. The data was made publicly available at [www.alzforum.org/exomes/hex](http://www.alzforum.org/exomes/hex).

##### 1.1.13 PERADES

The PERADES sample (Defining Genetic, Polygenic and Environmental Risk for Alzheimer’s Disease) comprises individuals with Alzheimer’s disease (AD) and healthy controls recruited across UK, Italy and Spain. The majority of the individuals are from the UK (n=4095 with samples recruited in Cardiff: n=2405), while the rest (n=841) were recruited in Spain and Italy. More specifically the recruitment centres were: MRC Centre for Neuropsychiatric Genetics and Genomics, Cardiff University, Cardiff, UK; Institute of Psychiatry, London, UK; University of Cambridge, Cambridge, UK; University of Southampton, Southampton, UK; University of Nottingham, Nottingham, UK; Catholic University of Rome, Rome, Italy; Santa Lucia Foundation, Rome, Italy; Instituto di Neurologia Policlinico Universitario, Rome, Italy; University of Milan, Milan, Italy; Laboratory of Gene Therapy, San Giovanni Rotondo, Italy; University of Perugia, Perugia, Italy; University of Cantabria and IDIVAL, Santander, Spain and the Regional Neurogenetic Centre (CRN), ASP Catanzaro, Lamezia Terme, Italy. The collection of the samples within the MRC Centre for Neuropsychiatric Genetics and Genomics, Cardiff University was through national recruitment through multiple channels, including specialist NHS services and clinics, research registers and Join Dementia Research (JDR) platform. The participants were assessed at home or in research clinics along with an informant, usually a spouse, family member or close friend, who provided information about and on behalf of the individual with dementia. Established measures were used to ascertain the disease severity: Bristol activities of daily living (BADL), Clinical Dementia Rating scale (CDR), Neuropsychiatric Inventory (NPI) and Global Deterioration Scale (GlDS). Individuals with dementia completed the Addenbrooke’s Cognitive Examination (ACE-r), Geriatric Depression Scale (GeDS) and National Adult Reading Test (NART) too. Control participants were recruited from GP surgeries and by means of self-referral (including existing studies and Joint Dementia Research platform). For all other recruitment, all AD cases met criteria for either probable (NINCDS-ADRDA, DSM-IV) or definite (CERAD) AD. All elderly controls were screened for dementia using the Mini Mental State Examination (MMSE) or ADAS-cog, were determined to be free from dementia at neuropathological examination or had a Braak score of 2.5 or lower. Control samples were chosen to match case samples for age, gender, ethnicity and country of origin. Informed consent was obtained for all study participants, and the relevant independent ethical committees approved study protocols. The whole exome sequencing (WES) was performed in-house at the MRC Centre for Neuropsychiatric Genetics and Genomics, Cardiff University. With the Nextera technology (Nextera Rapid Capture Exome v1.2), DNA was simultaneously fragmented and tagged with sequencing adapters in a single step. The enriched libraries were sequenced using the Illumina HiSeq 4000 (Illumina, USA) as paired-end 75 base reads according to manufacturer’s protocols.

##### 1.1.14 StEP-AD

The overall goals of the Stanford Extreme Phenotypes in AD (StEP AD) project are to identify and characterize novel genetic variants that promote resilience to AD pathology in the presence of the APOE4 allele or that drive pathogenesis in the absence of the APOE4 allele. Genomes are collected from several sources, some intramural and some extramural. Invariably, the cognitive assessment protocols for these different sources vary somewhat but all include APOE genotyping, extensive neuropsychological testing, collection of one or more AD biomarkers, and consensus adjudication. Genomes were sequenced for subjects in the following three categories: (1) Protected APOE4 carriers that have the APOE3/4 genotype, are at least 80 years old, and have normal cognition. If additional follow-up is expected we will accept subjects as young as 77; (2) Super-protected APOE4 carriers that have the APOE4/4 genotype, are at least 70 years old, and have normal cognition (if additional follow-up is expected subjects as young as 67 will be accepted); (3) APOE4-negative, early-onset cases that have the APOE2/2, 2/3, or 3/3 genotype and are diagnosed with probable AD before age 65. Most are also negative for known PSEN1, PSEN2 or APP mutations.

##### 1.1.15 Knight-ADRC

The samples Samples from the Charles F. and Joanne Knight Alzheimer’s Disease Research Center (Knight ADRC) were recruited at Washington University School of Medicine (WUSM) in Saint Louis, MO (USA). (REF). All the cases received a diagnosis of dementia of the Alzheimer's type (DAT), using criteria equivalent to the National Institute of Neurological and Communication Disorders and Stroke-Alzheimer's Disease and Related Disorders Association for probable AD^3,25^. Cognitively normal participants received the same assessment as the cases, and were deemed nondemented. Prior written consent, participants are genotyped for APOE4 allele and screened for known mutation in APP, PSEN1, PSEN2, MAPT, GRN, or C9orf72 by the Clinical and Genetics Core of the Knight ADRC. The approval number for the Knight ADRC Genetics Core family studies is 201104178.

##### 1.1.16 UCSF/NYGC/UAB

Studies in the UCSF/NYGC/UAB dataset were described previously^26^. Cases were selected from the University of California, San Francisco (UCSF) Memory and Aging Center with an intentional selection of early-onset cases to maximize the likelihood of identifying genetic contributors, along with healthy older adult controls (a total of 664 cases and 102 controls). All UCSF cases and controls were clinically assessed during an in-person visit to the UCSF Memory and Aging Center (MAC) that included a neurological exam, cognitive assessment, and medical history. Each participant’s study partner (i.e., spouse or close friend) was also interviewed regarding functional abilities. A multidisciplinary team composed of a neurologist, neuropsychologist, and nurse then established clinical diagnoses for cases according to consensus criteria. This cohort was intentionally depleted of cases with known Mendelian variants associated with neurodegenerative diseases. A small number of samples (19 cases and 21 controls) were obtained from the University of Alabama at Birmingham (UAB) from an expert clinician who employed the same diagnostic procedures.

##### 1.1.17 UCL-DRC EOAD

University College London Dementia Research Centre (UCL-DRC) early-onset Alzheimer’s disease cohort included patients seen at the Cognitive Disorders Clinics at The National Hospital for Neurology and Neurosurgery (Queen Square), or affiliated hospitals. Individuals were assessed clinically and diagnosed as having probable Alzheimer’s disease based on contemporary clinical criteria in use at the time, including imaging and neuropsychological testing where appropriate. All individuals consented for genetic testing and had causative mutations for Alzheimer’s disease (*PSEN1, PSEN2, APP*) and prion disease (*PRNP*) excluded prior to entry into this study.

##### 1.1.18  ADSP

**ADSP:** Cases and controls were selected from over 30,000 non-Hispanic Caucasian subjects from multiple cohorts described in detail elsewhere^27^. All controls were greater than 60 years and were cognitively normal based on direct assessment. All cases met NINCDS-ADRDA criteria for possible, probably, or definite Alzheimer’s disease. All cases had a documented age-at-onset, and for those with pathologically conformed AD, an age-at death. APOE genotypes were available for all. Cases were selected to have a minimal AD risk based on sex, age and APOE genotype. Controls were selected as those with the least probability of converting to AD by age 85. Controls were older (86.1 years, SD = 5.2) than cases (76.0 years, SD = 9.2). The selection criteria and the rationale for study design are described elsewhere^28^. Eventually, 5,096 cases and 4,965 controls were selected for exome sequencing by this protocol, as well as 682 additional cases from multiplex families with a strong AD family history.

**ADSP extension and Augmentation phase:** Under funding provided by NHGRI, an additional 3,000 subjects were whole genome sequenced. This included 1,466 cases and 1,534 controls. Of these 1,000 each of Non-Hispanic White (NHW), Caribbean Hispanic (CH), and African American (AA) descent were sequenced. Of these a total of 739 autopsy samples were sequenced [568 cases (500 NHW cases and 68 AA cases) and 171 controls (164 NHW and 7 AA)]. The Case-Control and Enriched Case Study spans 24 cohorts provided by the Alzheimer’s Disease Genetics Consortium (ADGC) and the Cohorts for Heart and Aging Research in Genomic Epidemiology (CHARGE) Consortium. The Augmentation Phase encompasses sequencing done under private and NIH funding by investigators who are not members of the ADSP. The investigators for these studies have agreed to share their GWAS, WGS and WES data with the ADSP. Private funding has been provided by industry and anonymous donors. Under the NIA AD Genetics Sharing Policy and the NIAGADS Data Distribution Agreement, individual NIA funded investigators studying the genetics and the genomics of AD provide their data to NIAGADS.

**Alzheimer’s Disease Neuroimaging Initiative (ADNI):** A public-private partnership, the purpose of ADNI is to develop a multisite, longitudinal, prospective, naturalistic study of normal cognitive aging, mild cognitive impairment (MCI), and early Alzheimer's disease as a public domain research resource to facilitate the scientific evaluation of neuroimaging and other biomarkers for the onset and progression of MCI and Alzheimer's disease. In 2017, ADNI geneticists began collaborations with the ADSP. Whole genome sequence data on 809 ADNI subjects (cases, mild cognitive impairment, and controls) have been harmonized using the ADSP pipeline. Data used in the preparation of this article were obtained from the Alzheimer’s Disease Neuroimaging Initiative (ADNI) database (adni.loni.usc.edu). The ADNI was launched in 2003 as a public-private partnership, led by Principal Investigator Michael W. Weiner, MD. The primary goal of ADNI has been to test whether serial magnetic resonance imaging (MRI), positron emission tomography (PET), other biological markers, and clinical and neuropsychological assessment can be combined to measure the progression of mild cognitive impairment (MCI) and early Alzheimer’s disease (AD). For up-to-date information, see [www.adni-info.org](http://www.adni-info.org).

##### 1.1.19 Variant calling

All contributing datasets were sequenced using a paired-end Illumina platform, but different exome capture kits were used, and a subset of the sample was sequenced using whole genome sequencing. For detailed information on variant and sample quality control and calling methods, please see Holstege and Hulsman et al., 2022^1^

#### 1.2 Effects of non-HPV and non-PTV rare SORL1 variants

##### 1.2.1 Effects of rare SORL1 variants with moderate priority: MPVs

MPVs have a relevant position in a functional domain as indicated by DMDM, but for which associated damagingness is currently unclear and requires more evidence (accompanying manuscript^29^). We identified 65 unique MPVs, carried by 80 AD cases and 61 controls. In aggregate, the risk of carrying an MPV associates with a 1.5-fold increased risk of AD (95%CI 1.1 - 2.1), and concentrates on the LOAD stage (OR=1.7, 95%CI 1.2 - 2.4, p=1.2x10^-1^). Compared to SORL1 WT patients, carriers of a MPV have an expedited AAO at later ages (**Fig 4A, Table 3 in manuscript**). MPVs with REVEL>50 associated with a 1.9-fold aggregated increased risk of (95%CI 1.3-3.0), p=2.2x10^-3^, indicating that several, but not all MPVs are functionally relevant. We identified 6 MPVs that affected the hydrophobic core of the YWTD domain (res 754-1013) at domain positions 6, 8, 15 and 42, carried by 6 cases and three controls. Furthermore, 2 patients and 3 controls carried the p.N924S substitution, affecting YWTD-domain at position 4 (part of the SBiN-motif) of the 5^th^ blade. In the CR-cluster (aa 1075-1550), we identified ten unique variants affecting the partly conserved glycine at position 38 which occurs in eight of the eleven CR-domains in SORL1, which were carried by 26 cases and 15 controls that affected in CR domains 5, 6, 9, 10, or 11, and in aggregate associated with a two-fold increased risk (OR = 2.0; 95%CI 1.06 – 3.8; p = 0.040). Of these, variant p.G1536S, was carried by 10 cases and 5 controls, contributing substantially to the aggregate effect of MPVs on AD risk. We further identified one AD patient who carried p.S1148R, which affects a domain-stabilizing serine at domain position 46, as part of an ‘Asx-turn’. Other MPVs in the CR domain were the fingerprint at residue 39, which we observed in 5 AD cases and 3 controls. We identified 14 MPVs in the 3Fn domains (aa 1551-2121): which were carried by 7 cases and 10 controls. Lastly, in the tail domain (aa 2161-2214), we observed 4 variants affecting the conserved FANSHY motif, which is essential for SORL1 binding to the retromer core complex: three cases carried respectively p.A2173T, p.S2175R (11:121498424:C>A), and p.H2176R. A fourth variant, p.S2175R (11:121498424:C>G) was carried by 10 cases and 11 controls. Lastly, two AD cases carried the p.D2207G variant in the GGA-binding motif of the SORL1 tail, involved in binding the adapter protein AP1. Overall, more research is necessary to understand the effect of specific tail-motif substitutions in the FANSHY or the GGA-binding motifs on AD risk.

##### 1.2.2 Effects of rare SORL1 variants with low and no priority: LPVs and NPVs

We identified 72 LPVs, which were carried by 85 AD cases with median AAO of 70 years (95%CI: 67-75) and 85 non-demented controls. Carrying an LPV does not associate with an increased risk of AD (OR=1.2, 95%CI (0.9 – 1.6; p=1) (**Table 3 in manuscript**). Similarly, we identified 267 NPVs, which were carried by 300 AD cases with a median AAO of 73 years, (95%CI: 71-74) and 312 controls. Carrying an NPV also does not associate with increased risk of AD (OR=1.1; 95%CI 0.9 - 1.3, p=1) (**Table 3**). Furthermore, a survival analysis indicated that carrying an LPV or an NPV does not lead to a significantly expedited AAO of AD relative to carriers of WT *SORL1* (**Fig 4A, Table 3 in manuscript**).

##### 1.2.3 Association of variants with MAF>0.05% with AD

A total of 8,578 individuals in our sample (21%) carried at least one of the 52 variants with MAF>0.05%, 18 of which were common enough for imputation in the latest AD GWAS^30^, allowing the identification of variant specific effects (**Table S2**). The most common coding *SORL1* variant, the p.A528T substitution (rs2298813, CADD score 25.5, REVEL score 0.112, carried by 3.6% of the individuals in the sample), associates with a 1.11-fold increased AD risk (p=5.79x10^-8^). We find no evidence for an effect on AD by the p.E270K substitution, carried by 1.9% of all individuals in the sample and that also affects the VPS10p domain: (rs117260922, CADD score 31, REVEL score 0.31, GWAS OR=1.02; p=5.89x10^-1^). The AAO of carriers from p.A528T and p.E270K variant carriers fully overlaps with carriers of WT *SORL1*. The p.D2065V substitution in the 3Fn domain (rs140327834, CADD score 28.4, REVEL score 0.568, carried by 0.46% of the sample), is associated with a 1.36-fold increased risk of AD in GWAS (p=1.61x10^-6^), and we observed a slightly expedited AAO for carriers. Our analysis provides no evidence that any of the other non-rare variants in our sample associates with altered risk of AD (**Fig 4C, Table S2)**.

#### 1.3 Comparison of DMDM with REVEL score and AlphaMissense.

Since the DMDM prioritization scheme could not be applied to the VPS10p domain, we relied on REVEL>0.5 to determine HPVs in the VPS10p domain. However, the AlphaMissense algorithm was released during our analyses, which led us to compare the performances between both algorithms when applied to all 1,103 carriers of the 511 unique rare coding SORL1 (MAF <0.05%) identified in our dataset. To investigate whether prioritization of the 107 *SORL1* variants by the manual DMDM approach outperformed prioritization by the in-silico by the REVEL algorithm^31^ (score-range: 0-1) and the AlphaMissense algorithm^32^ (score-range: 0-1), we compared its outcome in terms of effects on AD risk. Of the 107 HPVs, the REVEL score was not available for one variant and was therefore excluded, such that analyses were performed in comparison with 172 carriers of 106 HPV variants. For both the AlphaMissense and the REVEL algorithms, the effect of selected *SORL1* variants on AD risk increased with higher thresholds; compared to non-*SORL1*-variant carriers, variants with the highest REVEL scores associated with a 3.9-fold increased AD risk (95%CI:1.9-8.0), and variants with the highest AlphaMissense scores associated with 3.4-fold increased AD risk (95%CI: 2.1-5.5). In comparison, HPVs selected with the manual DMDM associated with a 6.1-fold AD-risk (95%CI: 4.1 – 9.0, p=5.3x10^-24^), indicating that the manual DMDM greatly outperformed the in silico-algorithms **(Fig S5-A)**. The number of unique rare variants that passed increasing thresholds declined similarly for each algorithm **(Fig S5-B)**. The variant prioritization scores generated by REVEL or AlphaMissense do not fully correlate (r = 0.52; 95%CI 0.48 - 0.56; p < 2.2x10^-16^), as many variants with REVEL<0.75 had low AlphaMissense scores, which indicates a differential reliance on variant-features between algorithms **(Fig S5-C)**. Notably, the scores of the variants in the VPS10p domain have correlated better (r = 0.80; 95%CI 0.75 - 0.84; p < 2.2x10^-16^), suggesting that these variants may have features that are weighted strongly by both algorithms. Nevertheless, several variants with REVEL scores <0.5 had AlphaMissense score >0.5, of which the majority affected the VPS10p domain. These variants were consequentially not annotated as HPVs, as we used the REVEL score >0.5 to select 27 VPS10p HPVs which, in aggregate, associated with an 8.8-fold increased risk of AD (95%CI 3.5- 22.3; p=3.4 x10^-7^). To investigate whether REVEL or AlphaMissense algorithms yielded the most damaging p.VPS10p variants, we compared the effect sizes. The 52 VPS10p variants selected with AlphaMissense score >0.5 associated with an aggregate 3.5-fold increased risk of AD (95%CI 2.2-5.7; p=8x10^-8^), while the 24 variants with REVEL>0.5 associated with an aggregate 6.9-fold increased risk of AD (95%CI 2.4-20.0; p=2.7x10^-5^). Furthermore, it seems that HPVs excluding those in VPS10p (which were preselected using REVEL) overlap more strongly with high REVEL scores than with high AlphaMissense scores. Together, our findings suggest that for our purpose of *SORL1* missense variant prioritization, using the REVEL algorithm might be preferred over the AlphaMissense algorithm, but that the manual DMDM outperforms both algorithms. This, in part, may explain the limited correlation between AlphaMissense and REVEL scores of the 106 HPVs: R=0.29 (95%CI 0.14 - 0.42; p = 1.3x10^-4^). Finally, we performed a Receiver Operating Characteristic (ROC) Analysis indicated a marginal performance improvement for the AlphaMissense algorithm over the REVEL algorithm, using a combination of both algorithms (by testing the sum of both scores) slightly surpasses individual performance **(Fig S5-D**).

### 2 Supplementary Figures and Tables

#### 2.1 Supplementary Figures

##### 2.1.1 Fig S1. PCA population

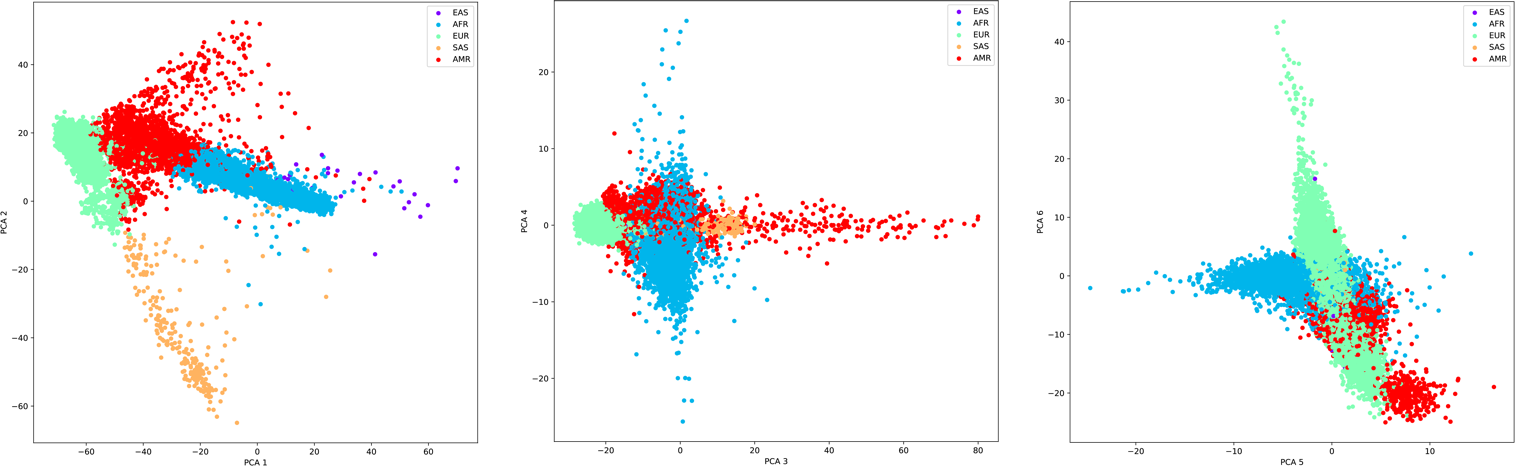

**Principal Component Analysis.** PCA1 vs. PCA2, PCA3 vs. PCA4, PCA5 vs. PCA6 were plotted across all samples for which whole exome sequencing was available, colored according to population group by 1000G (AFR = African, AMR = Admixed American, EAS = East Asian, EUR = European, SAS = South Asian). Note: Samples from Knight-ADRC and StEP-AD cohorts were not included here, as they were extracts of the coding sequences of the *SORL1*, *TREM2*, *ABCA7*, *ATP8B4*, *ABCA1*, *ADAM10*, *RIN3*, *CLU*, *ZCWPW1*, *ACE, and CBX3* genes as described previously^1^, and based on a separate PCA on these extracts, samples were annotated as EUR as we did not identify population outliers.

##### 2.1.2 Fig S2: The effects of MPVs, LPVs and NPVs in context of APOE.

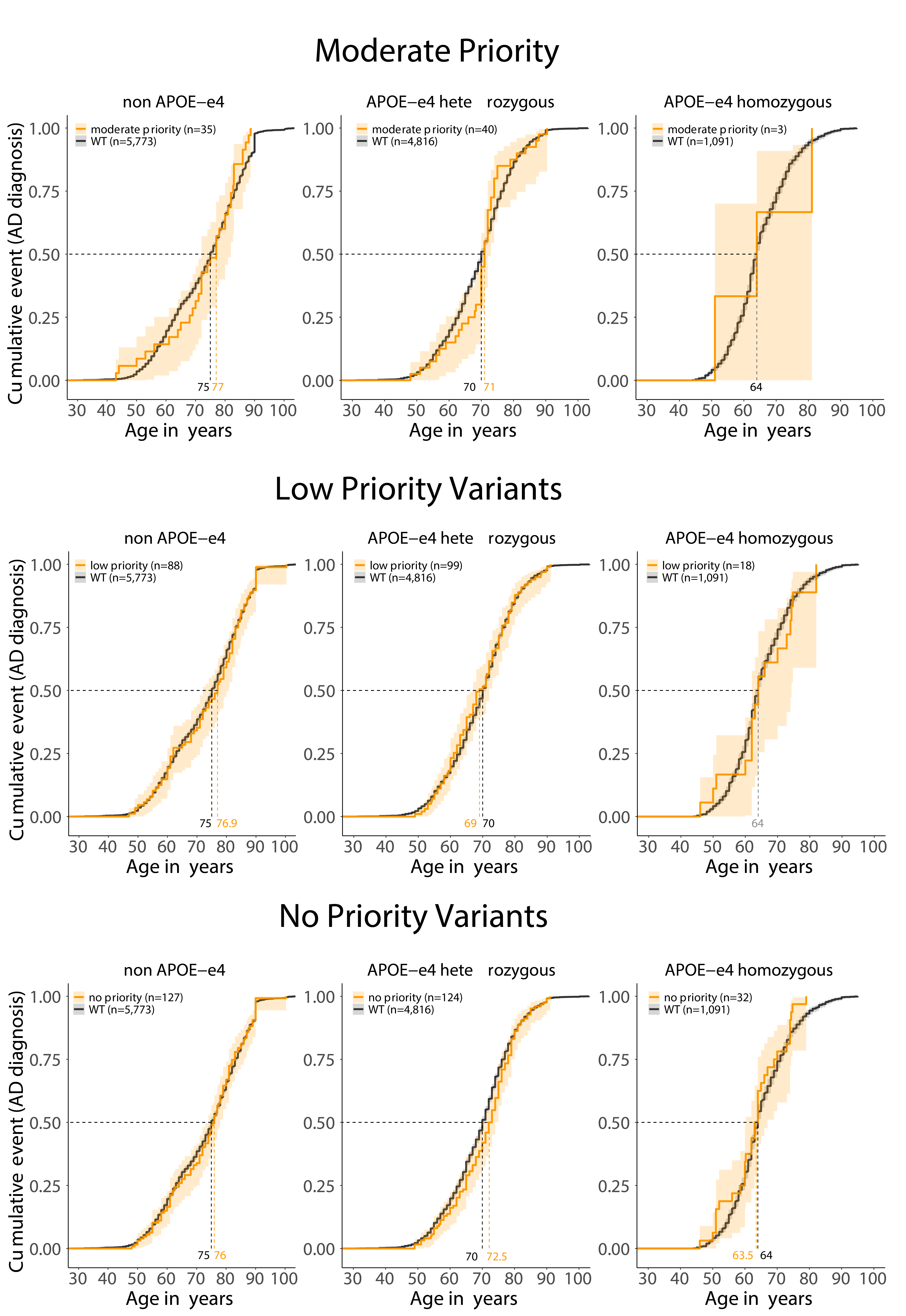

**A.** Age at onset of Moderate-priority variants relative to SORL1 WT carriers. The orange ages indicate at what age respectively 50% of the variant carriers had Alzheimer’s Disease, the black ages indicate at the age at which 50% of the SORL1 WT carriers developed AD. B. Low-priority variants in context of APOE genotype. C. No-priority variants in context of APOE genotype.

##### 2.1.3 Fig S3: Age at onset for specific variant carriers per priority group in context of APOE-genotype.

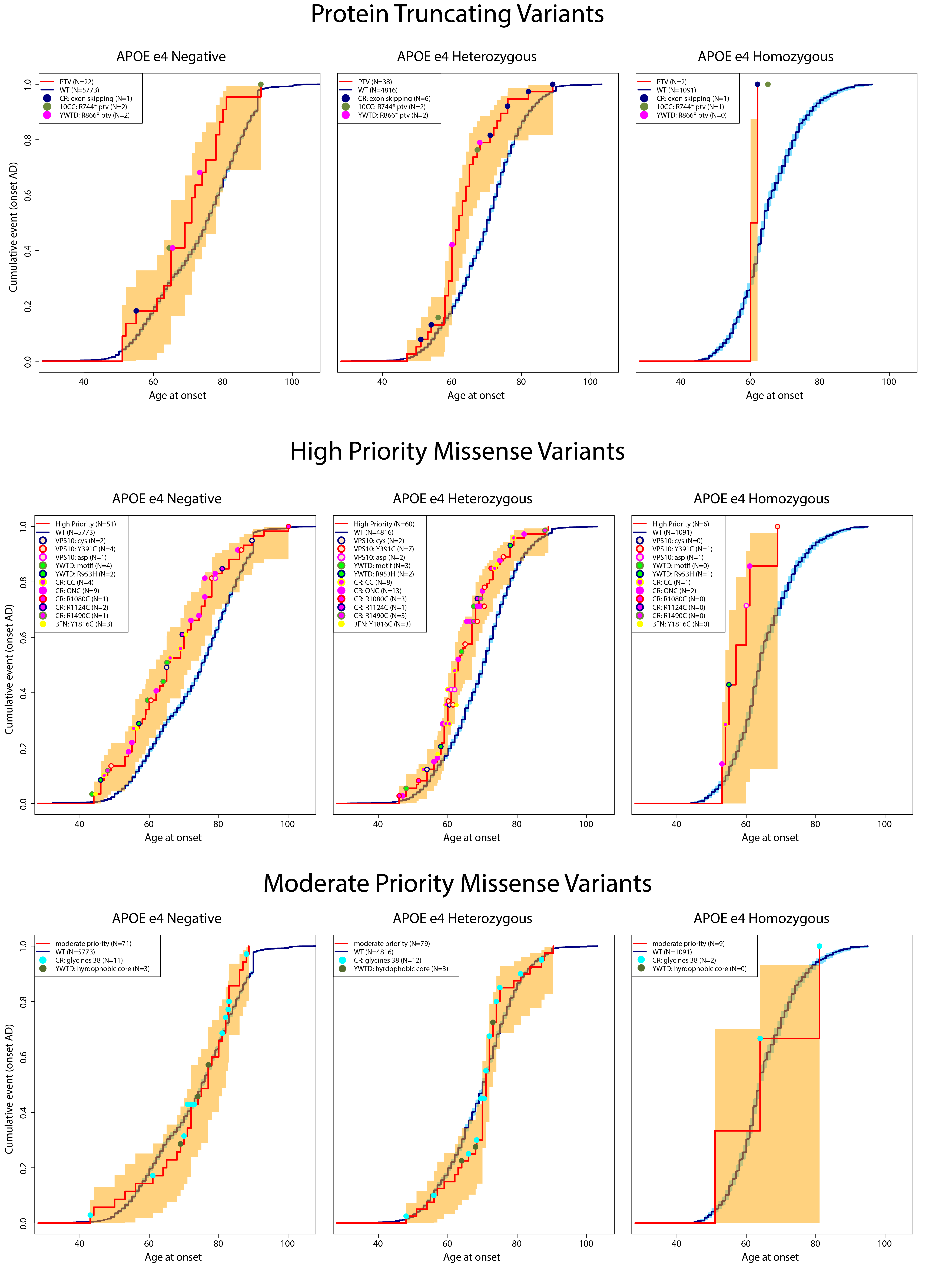

Age at onset of individual carriers with a specific variant (colored dot), in context of their *APOE*-genotype and priority group.

##### 2.2.1 Figure S4. Age at onset annotated by APOE genotype.

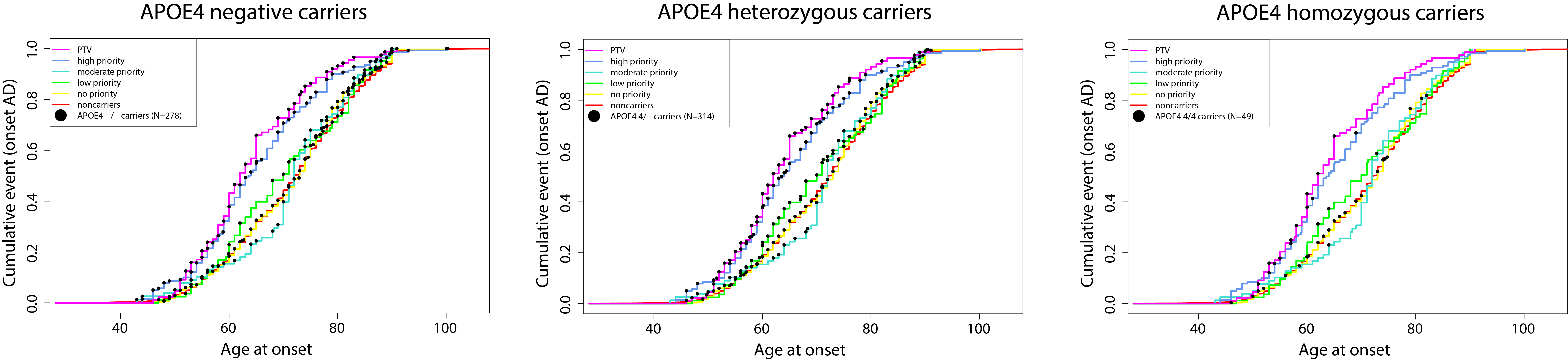

The age at onset of individual carriers with a specific APOE genotype (black dot), on prioritized categories.

##### 2.2.2 Figure S5: Comparison of DMDM with REVEL score and AlphaMissense

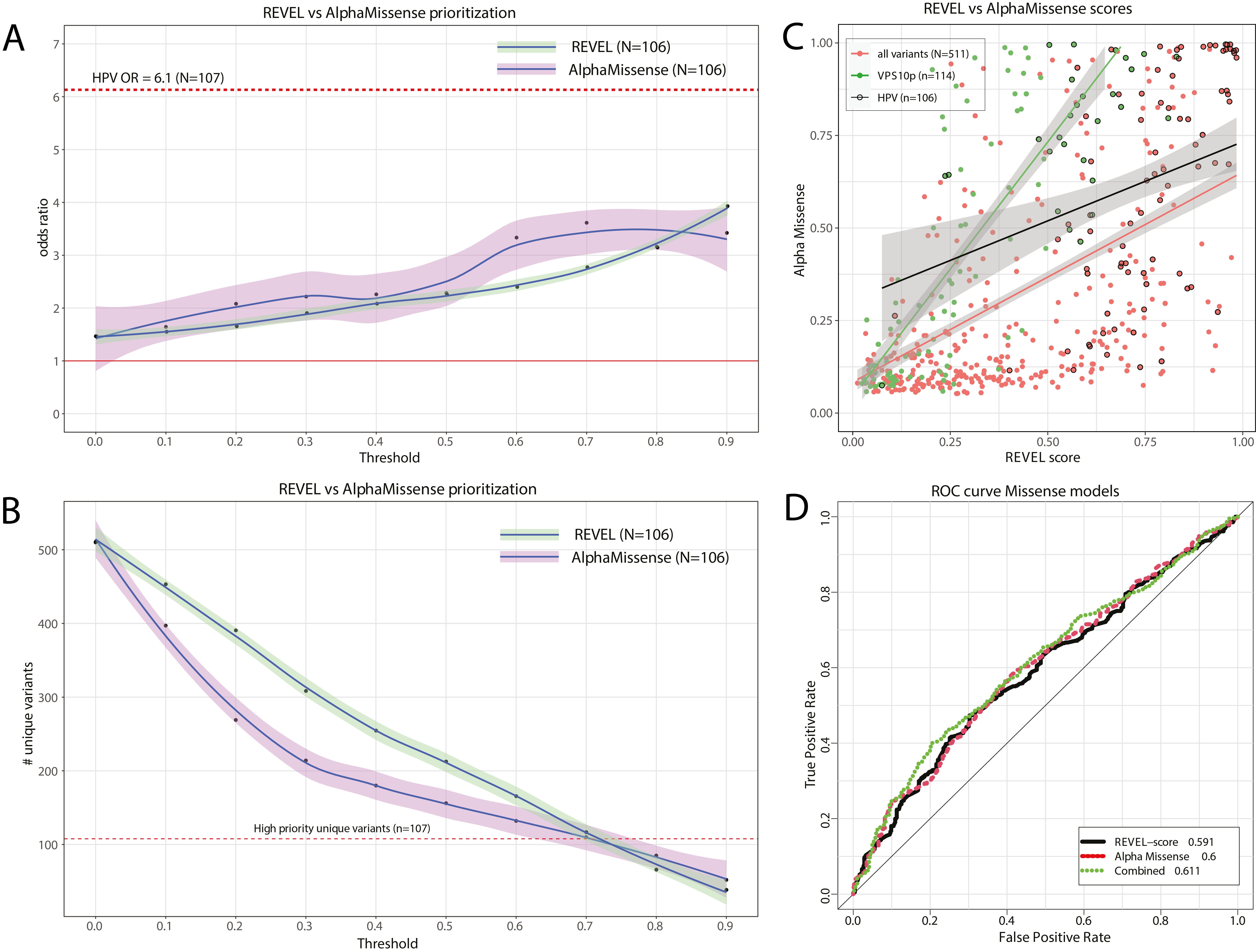

Analyses were performed with 172 carriers of 106 HPV variants, as the REVEL score was unavailable for one of the 107 HPVs. **A)** The increased risk of AD (odds ratio, y-axis), for carriers of variants with a REVEL or AlphaMissense threshold higher than a given threshold (x-axis). For comparison, the red dotted line indicates the OR in the HPV group. **B)** The number of unique variants (y-axis) with a REVEL or AlphaMissense score above a given threshold (x-axis); the red dotted line demonstrates the number of unique variants found in the high-priority category. **C)** Dotplot comparing REVEL scores with AlphaMissense scores for each variant considered in the study, annotated by variants occurring in the VPS10p domain, and variants considered HPVs. **D)** Receiver Operating Characteristic (ROC) Analysis in which the true positive and false positive rate for every unique threshold is calculated for all 106 variants for which REVEL score AND AlphaMissense scores were known. AD patients who carry a variant with an AlphaMissense score or REVEL score >X is regarded a ‘true positive’ while controls who carry a *SORL1* variant with an AlphaMissense score or REVEL score >X is regarded a ‘false positive’.

#### 2.3 Supplemental Tables:

The list of variants considered in the burden-analysis are available for download as spreadsheets in the Supplementary Data file.

##### 2.3.1 Table S1. XLS FILE: Rare variants (MAF < 0.05%)

##### 2.3.2 Table S2. XLS FILE: Non-rare variants (MAF > 0.05%)

##### 2.3.3 Table S3. XLS FILE: All excluded variants

| **SORLA motifs** | **domain pos** | **Tolerated**  **Substitutions** | **SORLA residues (aa number per subdomain)** |
| --- | --- | --- | --- |
| **VPS10p** |  |  |  |
| L2 Cys lost | Conserved disulfides | none | C^467^, C^473^ |
| L1 & L2 Cys gained | Disturb disulfide | NA | L1: 391 – 411; L2: 457 – 493; L1 & L2 protrusions |
| Asp-box specific variants in domain positions: with REVEL>0.5 | 42, 44, 46, 48,49 | NA | (β1) S^138^, D^140^, G^142^, S^144^, F^145^, (β2) T^190^, D^192^, NA, T^196^, NA (β3) S^234^, D^236^, G^238^, T^240^, W^241^, (β4) S^280^, D^282^, NA, S^286^, NA, (β5) S^329^, NA, NA, NA, NA, (β6) S^375^, NA, G^379^, NA, F^382^, (β7) T^443^, D^445^, G^447^, T^449^, W^450^, (β8) S^523^, NA, G^527^, NA, W^530^, (β9) S^564^, N^566^, G^568^, T^570^, W^571^  Positions are likely most pathogenic when affecting a ‘conserved’ residue |
| Variants with REVEL>0.5 | Random | NA | Positions are likely most pathogenic when affecting a ‘conserved’ residue |
| **10CC** |  |  |  |
| ONC (Cys loss) | Conserved disulfides | none | (CCa) C^625^, C^643^, C^660^, C^675^, (CCb) C^677^, C^684^, C^699^, C^716^, C^736^, C^752^ |
| ONC (Cys gained) | Disturb disulfide | NA | Random |
| **YWTD** |  |  |  |
| YWTD-motif | 16, 17, 18, 19 | none | (β2) Y^803^, W^804^, S^805^, D^806^; (β3) Y^847^, W^848^, NA, D^850^; (β4) F^891^, W^892^, T^893^, D^894^; (β5) Y^934^, W^935^, T^936^, D^937^; (β6) Y^974^, W^975^, NA, D^977^ |
| Partly conserved Pro | 3 | none | (β4) P^878^, (β5) P^923^, (β6) P^963^ |
| Partly conserved Asp | 9 | none | (β2) D^796^, (β5) D^929^ |
| Highly conserved Ile | 27 | none | (β1) I^769^, (β2) I^812^, (β3) I^856^, (β4) I^902^, (β5) I^943^, (β6) I^983^ |
| Highly conserved Arg | 29 | none | (β1) R^771^, (β2) R^814^, (β3) NA, (β4) R^904^, (β5) R^945^, (β6) R^985^ |
| Highly conserved G | 35 | none | (β1) G^777^, (β2) G^819^, (β3) G^863^, (β4) G^909^, (β5) G^950^, (β6) G^991^ |
| Partly conserved R* | 38 | none | (β3) R^866^, (β5) R^953^; DMDM only when R changed |
| Cysteines Disulfide β2 | Likely disulfide | none | (β2) C^801^, (β2) C^816^ |
| **EGF** |  |  |  |
| ONC (Cys loss) | Conserved disulfides | none | C^1021^, C^1026^, C^1030^, C^1040^, C^1042^, C^1058^, C^1060^, C^1071^ |
| ONC (Cys gain) | Disturb disulfides | NA | random |
| **CR** |  |  |  |
| Calcium cage (D,D,D,E) | 37, 41, 47, 48 | none | (CR1) D^1098^, D^1102^, D^1108^, E^1109^; (CR2) D^1139^, D^1143^, D^1149^, E^1150^; (CR3) D^1178^, D^1182^, D^1188^, E^1189^; (CR4) D^1219^, D^1223^, D^1229^, E^1230^; (CR5) D^1257^, D^1261^, D^1267^, E^1268^; (CR6) D^1297^, Q^1301^, D^1307^, E^1308^; (CR7) D^1345^, D^1349^, D^1355^, E^1356^; (CR8) D^1389^, D^1393^, D^1399^, E^1400^; (CR9) D^1439^, D^1443^, D^1449^, E^1450^; (CR10) D^1492^, D^1496^, D^1502^, E^1503^; (CR11) D^1535^, D^1539^, D^1545^, E^1546^ |
| ONC (Cys loss) | 15, 23, 29, 36, 42, 55  Conserved disulfides | none | (CR1) C^1078^, C^1085^, C^1090^, C^1097^, C^1103^, C^1112^; (CR2) C^1117^, C^1125^, C^1131^, C^1138^, C^1144^, C^1153^; (CR3) C^1158^, C^1165^, C^1170^, C^1177^, C^1183^, C^1192^; (CR4) C^1199^, C^1206^, C^1211^, C^1218^, C^1224^, C^1235^; (CR5) C^1239^, C^1244^, C^1249^, C^1256^, C^1262^, C^1271^; (CR6) C^1275^, C^1283^, C^1289^, C^1296^, C^1302^, C^1315^; (CR7) C^1325^, C^1332^, C^1337^, C^1344^, C^1350^, C^1359^; (CR8) C^1368^, C^1376^, C^1381^, C^1388^, C^1394^, C^1403^; (CR9) C^1419^, C^1426^, C^1431^, C^1438^, C^1444^, C^1453^; (CR10) C^1471^, C^1478^, C^1484^, C^1491^, C^1497^, C^1506^; (CR11) C^1514^, C^1521^, C^1527^, C^1534^, C^1540^, C^1549^ |
| ONC (Cys gain) | Disturb disulfides | NA | random |
| Asx-turn (D) | 44 | none | (CR1) D^1105^, (CR2) D^1146^, (CR3) D^1185^, (CR4) D^1226^, (CR5) D^1264^, (CR6) D^1304^, (CR7) D^1352^, (CR8) D^1396^, (CR9) D^1446^, (CR10) D^1499^, (CR11) D^1542^ |
| **3Fn** |  |  |  |
| Partly conserved prolines | P’s at 6, 7, 28, 79 | none | (3FN1) NA, NA; P^1578^, P^1619^; (3FN2) P^1654^, NA; P^1676^, NA; (3FN3) P^1749^, P^1750^; NA, NA; (3FN4) P^1843^, P^1844^, P^1865^, NA; (3FN5) P^1934^, P^1935^, P^1955^, P^1998^; (3FN6) NA, P^2027^, NA, NA |
| W; B-strand | 25 | none | (3FN1) W^1575^, (3FN2) W^1673^, (3FN3) (L^1767^), (3FN4) W^1862^, (3FN5) W^1952^, (3FN6) W^2043^ |
| Partly conserved glycines | G’s at 36, 96 | none | (3FN2) G^1681^, (3FN2) G^1732^, (3FN4) G^1917^ |
| Y; C-strand | 41 | aromatic residues tolerated | (3FN1) Y^1588^, (3FN2) Y^1686^, (3FN3) Y^1778^, (3FN4) Y^1870^, (3FN5) Y^1965^, (3FN6) Y^2059^ |
| L; EF-loop (tyrosine corner) | 77 | none | (3FN1) L^1617^, (3FN2) L^1713^, (3FN3) L^1810^, (3FN4) NA, (3FN5) L^1996^, (3FN6) L^2087^ |
| Y; F-strand (tyrosine-corner) | 83 | none | (3FN1) Y^1623^, (3FN2) Y^1719^, (3FN3) Y^1816^, (3FN4) Y^1905^, (3FN5) Y^2002^, (3FN6) Y^2093^ |
| Arg hotspot | 88: | none | (3FN4) R^1910^ Based on Arg substitutions in DMDM pathogenic |
| Cysteines 1^st^ domain | 39, 91, disulfide | none | (3FN1) C^1586^, (3FN1) C^1631^ |
| Cysteines 6^th^ domain | 91, 98: disulfide | none | (3FN6) C^2101^, (3FN6) C^2108^ |

##### 2.3.4 Table S4: List of high-priority variants based on DMDM analysis and sequence conservation.

Prioritization of positions likely to harbor pathogenic mutations based on either domain sequence conservation or because the DMDM analysis. Analysis of enrichment: number of observations in sample (cases and controls) per possible positions. NA: in the specific domain, the prioritized variant does not exist. * When Arg at position 38 in the YWTD-domain of LDLR, LRP4 or LRP5 are substituted by other residues, this leads to autosomal dominant inherited forms of diseases. SORL1 only has a Arg at position 38 in β3 and β5: R^866^ and R^953^.

##### 2.3.5  Table S5: Moderate-priority variants

| **SORLA motifs** | **domain pos** | **Tolerated**  **Substitutions** | **SORLA residue/aa no., per subdomain** |
| --- | --- | --- | --- |
| **VPS10p** |  |  |  |
| RGD (in ProP) | NA | none | R^63^, G^64^, D^65^ |
| RRKR (in ProP) | NA | none | R^78^, R^79^, K^80^, R^81^ |
| Hydrophobic; A-strand | 5, 6 | Conservative (hydrophobic) substitutions likely tolerated | (β1) V^106^, V^107^, (β2) F^167^, Y^168^, (β3) L^209^, L^210^, (β4) NA, F^251^, (β5) F^300^, NA, (β6) Y^349^, Y^350^, (β7) F^414^, NA, (β8) L^495^, NA, (β9) Y^539^, Y^540^, (β10) V^583^, Y^584^ |
| Hydrophobic; B-strand | 19, 20, 21 | Conservative (hydrophobic) substitutions likely tolerated | (β1) I^117^, V^118^, A^119^, (β2) Y^177^, I^178^, F^179^, (β3) L^218^, L^219^, L^220^, (β4) I^264^, Y^265^, I^266^, (β5) Y^306^, M^307^, F^308^, (β6) V^359^, F^360^, V^361^, (β7) Y^424^, I^425^, A^426^, (β8) I^504^, I^505^, A^506^, (β9) I^548^, I^549^, NA, (β10) V^596^, F^597^, NA |
| Hydrophobic; C-strand | 39, 40, 41 | Conservative (hydrophobic) substitutions likely tolerated | (β1) V^135^, Y^136^, V^137^, (β2) L^187^, W^188^, I^189^, (β3) L^231^, W^232^, NA, (β4) V^277^, F^278^, NA, (β5) L^326^, W^327^, V^328^, (β6) L^372^, Y^373^, I^374^, NA, V^441^, I^442^, (β7) V^520^, Y^521^, I^522^, (β8) L^561^, NA, Y^563^, (β9) L^613^, NA, V^615^ |
| **YWTD** |  |  |  |
| hydrophobic | 6, 8, 15 | Conservative (hydrophobic) substitutions likely tolerated | (β2) L^793^, F^795^, L^802^; (β3) L^837^, F^839^, L^846^; (β4) L^881^, L^883^, M^890^; (β5) I^926^, V^928^, I^933^; (β6) I^966^, V^968^, I^973^; (β1) M^1007^, I^1009^, NA |
| hydrophobic | 41, 42 | Conservative (hydrophobic) substitutions likely tolerated | (β1) NA, L^782^; (β2) V^825^, I^826^; (β3) I^869^, V^870^; (β4) L^915^, V^916^; (β5) I^956^, L^957^; (β6) I^996^, L^997^ |
| Highly conserved L | 47 | none | (β1) L^787^, (β2) L^831^, (β3) L^875^, (β4) V^920^, (β5) L^960^, (β6) L^1001^ |
| SBIN, NXI,  Ligand binding? | 4, 20 blades β4, β5, β6 | uncertain | (β4) R^879^, (β4) W^895^, (β5) N^924^, (β6) Y^964^, (β6) W^978^ |
| **CR** |  |  |  |
| Asx-turn (S) | 46 | none | (CR1) S^1107^, (CR2) S^1148^, (CR3) S^1187^, (CR4) S^1228^, (CR5) S^1266^, (CR6) S^1306^, (CR7) S^1354^, (CR8) S^1398^, (CR9) S^1448^, (CR10) NA, (CR11) S^1544^ |
| Fingerprints, Involved in ligand binding | 34, 39 | uncertain | (CR1) W^1095^, D^1100^; (CR2) Y^1136^, E^1141^; (CR3) W^1175^, D^1180^; (CR4) W^1216^, D^1221^; (CR5) K^1254^, L^1259^; (CR6) M^1294^, I^1299^; (CR7) W^1342^, M^1347^; (CR8) W^1386^, E^1391^; (CR9) W^1436^, Y^1441^; (CR10) K^1489^, H^1494^; (CR11) E^1532^, F^1537^ |
| Phe-Ile hyd core,  Strong conservation | 21, 30 | Uncertain, (few variants in DMDM analysis) | (CR1) Y^1083^, I^1091^; (CR2) F^1123^, I^1132^; (CR3) Y^1163^, I^1171^; (CR4) F^1204^, I^1212^; (CR5) F^1242^, I^1250^; (CR6) F^1281^, L^1290^; (CR7) F^1330^, I^1338^; (CR8) F^1374^, I^1382^; (CR9) Y^1424^, V^1432^; (CR10) F^1476^, I^1485^; (CR11) F^1519^, I^1528^ |
| Partly conserved glycines, (Few variants in DMDM analysis) | 27, 38 | uncertain | (CR1) G^1088^, NA; (CR2) G^1129^, NA; (CR3) G^1168^, G^1179^; (CR4) G^1209^, G^1220^; (CR5) G^1247^, G^1258^; (CR6) NA, G^1298^; (CR7) G^1335^, G^1346^; (CR8) G^1379^, NA; (CR9) G^1429^, G^1440^; (CR10) NA, G^1493^; (CR11) NA, G^1536^ |
| **3Fn** |  |  |  |
| Hydrophobic | 11, 13 | Conservative (hydrophobic) substitutions likely tolerated | (3FN1) L^1561^, W^1563^; (3FN2) L^1659^, L^1661^; (3FN3) I^1753^, I^1755^; (3FN4) L^1848^, A^1850^; (3FN5) L^1938^, V^1940^; (3FN6) L^2030^, I^2032^ |
| Hydrophobics in B-strand | 21, 23 | Conservative (hydrophobic) substitutions likely tolerated | (3FN1) V^1571^, L^1573^; (3FN2) I^1669^, NA; (3FN3) L^1763^, F^1765^; (3FN4) V^1858^, C^1860^; (3FN5) V^1948^, I^1950^; (3FN6) V^2039^, L^2041^ |
| Hydrophobics in C-strand | 43, 45 | Conservative (hydrophobic) substitutions tolerated | (3FN1) V^1590^, Y^1592^; (3FN2) V^1688^, Y^1690^; (3FN3) V^1780^, L^1782^; (3FN4) I^1872^, Y^1874^; (3FN5) V^1967^, V^1969^; (3FN6) I^2061^, M^2063^ |
| Partly conserved glycines | 94 | uncertain | (3FN2) G^1730^, (3FN3) G^1827^, (3FN6) G^2104^ |
| Hydrophobics in F-strand | 85, 87, 89 | Conservative (hydrophobic) substitutions tolerated | (3FN1) V^1625^, V^1627^, V^1629^; (3FN2) V^1721^, V^1723^, A^1725^; (3FN3) I^1818^, A^1820^, A^1822^; (3FN4) F^1907^, V^1909^, V^1911^; (3FN5) I^2004^, V^2006^, L^2008^; (3FN6) F^2095^, V^2097^, A^2099^ |
| Hydrophobics in E-strand | 72, 74 | Conservative (hydrophobic) substitutions tolerated | (3FN1) NA, L^1614^; (3FN2) NA, I^1710^; (3FN3) NA, V^1807^; (3FN4) V^1897^, V^1899^; (3FN5) Y^1991^, L^1993^; (3FN6) F^2082^, I^2084^ |
| Trp-ladders |  | uncertain | (3FN1) R^1593^, W^1600^, K^1626^, H^1636^; (3FN2) E^1690^, W^1698^, R^1722^, W^1734^ |
| **Tail** |  |  |  |
| FANSHY |  | uncertain | F^2172^, A^2173^, N^2174^, S^2175^, H^2176^, Y^2177^ |
| Acidic |  | uncertain | D^2190^, D^2191^, L^2192^, G^2193^, E^2194^, D^2195^, D^2196^, E^2197^, D^2198^ |
| GGA |  | uncertain | D^2207^, D^2208^, V^2209^, P^2210^, M^2211^, V^2212^, I^2213^, A^2214^ |

Rare variants (MAF<0.05%) in these positions are considered moderate-priority. Variants correspond with the grey residues in **Figure 3**. NA: the specific domain does not have the prioritized variant. Underlined: Note that we observed a moderate-priority variant leading to a W1563C substitution of a hydrophobic residue at position 13 in the 3Fn domain. This is a non-rare variant, with a non-neuro pop-max MAF of 6.7x10^-4^ (GnomAD) a REVEL score of 0.476 and a CADD score of 32. We observed no evidence for an effect on AD of this variant in the latest GWAS: OR=0.95 (95% CI 0.56-1.6], p=8.35x10^-1^.

##### 2.3.6 Table S6. Age at onset per variant carrier group relative to WT SORL1 carriers

| **SORL1 variant type** | **Median age at onset**  **95%CI on median** | **10%-90%**  **Inter percentile range** | **∆**  **median AAO vs WT  (95% CI; p value*)** | **∆**  **median AAO vs PTV  (95% CI, p value*)** |
| --- | --- | --- | --- | --- |
| **Fig 4a** |  |  |  |  |
| SORL1 WT | 72 (72 - 73) | 56-87 | NA | 10 (12 — 8); 2.7E-11 |
| PTV | 62 (60 - 65) | 52-78 | -10 (-12 — -8); 2.7E-11 | NA |
| High-priority missense | 64 (62 - 67) | 53-79 | -8 (-10 — -6); 5.3E-9 | 2 (2 – 2); 1 |
| Moderate-priority missense | 72 (71 - 74) | 54-86 | 0 (-1 — 1); 1 | 10 (11 – 9); 2.3E-3 |
| Low-priority missense | 70 (67 - 75) | 56-84.9 | -2 (-5 — 2); 1 | 8 (7 – 10); 6.9E-3 |
| No-priority missense | 73 (71 - 74) | 55-86 | 1 (-1 - 1); 1 | 11 (11 – 9); 3.3E-7 |
| **Fig 4b** |  |  |  |  |
| YWTD-motif | 64 (48 - NA) | 44-68 | -8 (-24 — NA); 2.6E-3 | 2 (-12 – NA); 1 |
| calcium cage | 60 (56 - NA) | 54-73 | -12 (-16 — NA); 7.7E-4 | -2 (-4 – NA); 1 |
| VPS10 REVEL>50 | 59.5 (56 - 78) | 46-83 | -12.5 (-16 —5); 1 | -2.5 (-4 – 13); 1 |
| ONC in CR domain | 68 (63 - 74.3) | 53-82 | -4 (-9 — 1.3); 1 | 6 (3 – 9.3); 1 |
| 10CC | 67 (63 - 73) | 57-78 | -5 (-9 — 0); 1 | 5 (3 – 8); 1 |
| Y391C VPS10p Loop1 | 67.5 (60.1 - NA) | 60-78 | -4.5 (-11.9 — NA); 1 | 5.5 (0.1 – NA); 1 |
| other HPV carriers | 61 (60 - 70) | 53-79 | -11 (-12 — -3); 9.4E-3 | -1 (0 – 5); 1 |
| **Fig 4C** |  |  |  |  |
| D2065V | 70 (68 - 72) | 53.1-85 | -2 (-4 — -1); 7.1E-1 | 8 (8 – 7); 9.7E-4 |
| E270K | 72 (71 - 73) | 56-87 | 0 (-1 — 0); 1 | 10 (11 – 8); 5.2E-9 |
| A528T | 71 (70 - 72) | 55-86 | -1 (-2 — -1); 1.7E-1 | 9 (10 – 7); 4.8E-8 |
| other MAF >0.05% | 74 (73 - 74) | 57-87 | 2 (1 — 1); 1 | 12 (13 – 9); 4.9E-13 |

The estimated median age at onset per priority group or specific variant carrier group and the difference between the age at onset of carriers of WT *SORL1*. For all the different variant groups, both the age at onset when 50% and 80% of the variant carriers developed AD is given. *P value was calculated using a log-rank test. See **Figure 4** for age distributions per variant group. ONC: odd numbered cysteines (gain or loss of a cysteine).

##### 2.3.7 Table S7. Differences in age at onset per priority category in context of APOE genotype.

| **SORL1 variant type** | **APOE-4 genotype** | **Median age at onset (95%CI)** | **10%-90%**  **Inter percentile range** | **∆ median AAO**  **v.s. WT  (95% CI); p value*** | **∆ median AAO**  **v.s. PTV  (95% CI); p value*** |
| --- | --- | --- | --- | --- | --- |
| **Fig 5** |  |  |  |  |  |
| WT | E4-NEG | 75 (75 - 76) | 56-89 | NA | 6 (10 — 2); 2.7E-2 |
|  | E4-HET | 70 (70 - 71) | 55-82 | NA | 9 (10 — 6); 4.0E-6 |
|  | E4-HOM | 64 (63 - 64) | 54-77 | NA | 4 (6 - NA); 9.2E-2 |
| PTV | E4-NEG | 69 (65 - 74) | 52-81 | -6 (-10 — -2); 2.7E-2 | NA |
|  | E4-HET | 61 (60 - 65) | 51-74 | -9 (-10 — -6); 4.0E-6 | NA |
|  | E4-HOM | 60 (57 - NA) | 53-65 | -4 (-6 - NA); 9.2E-2 | NA |
| HPV | E4-NEG | 66 (62 - 74.3) | 48-86 | -9 (-13 – -1.7); 1.7E-2 | -3 (-3 - 0.3); 1 |
|  | E4-HET | 63 (62 - 67) | 54-78 | -7 (-8 —-4); 1.6E-4 | 2 (2 - 2); 1 |
|  | E4-HOM | 58.5 (55 - NA) | 53-69 | -5.5 (-8 — NA); 2.9E-2 | -1.5 (-2 - NA); 1 |
| **Fig S2** |  |  |  |  |  |
| MPV | E4-NEG | 77 (72 - 82) | 53-86 | -2 (-3 — 6); 1 | 8 (7 - 8); 0.9 |
|  | E4-HET | 71 (70 - 73) | 56.5-82.4 | 1 (0 - 2); 1 | 10 (10 - 8); 2.7E-2 |
|  | E4-HOM | 64 (51 - NA) | 51-81.1 | 0 (-12 - NA); 1 | 4 (-6 - NA); 1 |
| LPV | E4-NEG | 78.2 (71 - 83) | 53-88 | 3.2 (4 - 7); 1 | 9.2 (6 - 9); 0.4 |
|  | E4-HET | 66 (63 - 71) | 56-81 | -4 (-7 - 0); 1 | 5 (3 - 6); 0.8 |
|  | E4-HOM | 72.8 (64 - NA) | 62-82 | -8.8 (1 - NA); 1 | 12.8 (7 - NA); 0.2 |
| NPV | E4-NEG | 76 (74 - 78) | 55-89 | 1 (1 - 2); 1 | 7 (9 - 4); 0.1 |
|  | E4-HET | 72.5 (70 - 74) | 57-82 | 2.5 (0 - 3); 1 | 11.5 (10 - 9); 1.5E-05 |
|  | E4-HOM | 63.5 (60.3 - 67) | 51-74 | -0.5 (-2.7 - 3); 1 | 3.5 (3.3 - NA); 0.7 |

The estimated median age at onset per priority group or specific variant carrier group, in context of its APOE-E4 genotype, and the difference between the age at onset of carriers of WT *SORL1*. For all the different variant groups, both the age at onset when 50% and 80% of the variant carriers developed AD is given. *P value was calculated using a log-rank test.

### 3 ACKNOWLEDGMENTS

#### 3.1 Study participants, personnel, and compute infrastructure

##### 3.1.1 Participants

The authors are grateful to the study participants, their family members, and the participating general practitioners, pharmacists and all laboratory personnel involved in blood collection, DNA isolation, and DNA biobanking.

##### 3.1.2 SURF supercomputer facility

The work in this manuscript was carried out on the Cartesius supercomputer, which is embedded in the Dutch national e-infrastructure with the support of SURF Cooperative. Computing hours were granted in 2016, 2017, 2018 and 2019 to H. Holstege by the Dutch Research Council (project name: ‘100plus’; project numbers 15318 and 17232).

#### 3.2 Study Cohorts

##### 3.2.1 ADES-FR

This study was funded by grants from the Clinical Research Hospital Program from the French Ministry of Health (GMAJ, PHRC, 2008/067), the CNR-MAJ, the JPND PERADES, Equipe FRM DEQ20170336711, and Fondation Alzheimer (ECASCAD study). This research was supported by the Laboratory of Excellence GENMED (Medical Genomics) grant no. ANR-10-LABX-0013 managed by the National Research Agency (ANR) part of the Investment for the Future program. This work was also supported by Foundation Alzheimer, the Institut Pasteur de Lille, Inserm, the Haut-de-France and Lille Métropole Communauté Urbaine council, and the French government's LABEX (laboratory of excellence program investment for the future) DISTALZ grant (Development of Innovative Strategies for a Transdisciplinary approach to Alzheimer's disease). The 3C Study supports are listed on the Study Website ([www.three-city-study.com](https://eur04.safelinks.protection.outlook.com/?url=http%3A%2F%2Fwww.three-city-study.com%2F&data=02|01||77be9bf957134a87b79c08d82c8120e6|68dfab1a11bb4cc6beb528d756984fb6|0|0|637308280402083619&sdata=rMMvVXe%2F8w6dqfHOemOhHS8tBaPy2XUzXGEQOPbVevY%3D&reserved=0)).

##### 3.2.2 AgeCoDe-UKBonn

The AgeCoDe cohort was funded in part by the German Federal Ministry of Education and Research (BMBF) (grants KNDD 01GI0710, 01GI0711, 01GI0712, 01GI0713, 01GI0714, 01GI0715, 01GI0716, 01ET1006B). Sequencing of AgeCoDe sample was in part funded by the German Research Foundation (DFG) grant RA 1971/6-1 to Alfredo Ramirez.

##### 3.2.3 Barcelona- SPIN

Support for Jordi Clarimon provided by Maratón RTVE (Spain).Support for Oriol Dols provided by the Association for Frontotemporal Degeneration (Clinical Research Postdoctoral Fellowship, AFTD).

##### 3.2.4 AC-EMC

Exome sequencing was funded by Alzheimer Nederland.

##### 3.2.5 ERF

The ERF study as a part of EUROSPAN (European Special Populations Research Network) was supported by European Commission FP6 STRP grant number 018947 (LSHG-CT-2006-01947) and also received funding from the European Community's Seventh Framework Programme (FP7/2007-2013)/grant agreement HEALTH-F4- 2007-201413 by the European Commission under the programme "Quality of Life and Management of the Living Resources" of 5th Framework Programme (no. QLG2-CT-2002- 01254). High-throughput analysis of the ERF data was supported by a joint grant from the Netherlands Organization for Scientific Research and the Russian Foundation for Basic Research (NWO-RFBR 047.017.043).

##### 3.2.6 Rotterdam Study

The generation and management of the exome sequencing data for the Rotterdam Study was executed by the Human Genotyping Facility of the Genetic Laboratory of the Department of Internal Medicine, Erasmus MC, the Netherlands. The Rotterdam Study is funded by Erasmus Medical Center and Erasmus University, Rotterdam, the Netherlands Organization for Health Research and Development (ZonMw), the Research Institute for Diseases in the Elderly (RIDE) (014-93-015; RIDE2), the Ministry of Education, Culture and Science, the Ministry for Health, Welfare and Sports, the European Commission (DG XII), and the municipality of Rotterdam. Genetic data sets are also supported by the Netherlands Organization of Scientific Research NWO Investments (175.010.2005.011, 911-03-012), the Genetic Laboratory of the Department of Internal Medicine, Erasmus MC, and the Netherlands Genomics Initiative (NGI)/Netherlands Organization for Scientific Research (NWO), the Netherlands Consortium for Healthy Aging (NCHA), project 050-060-810, and by a Complementation Project of the Biobanking and Biomolecular Research Infrastructure Netherlands (BBMRI-NL; www.bbmri.nl ; project number CP2010-41). We thank Mr. Pascal Arp, Ms. Mila Jhamai, Mr. and Marijn Verkerk, for their help in creating the RS-Exome Sequencing database.

##### 3.2.7 ADC-Amsterdam

We thank all study participants and all personnel involved in data collection for the contributing studies. Research of Alzheimer center Amsterdam is part of the neurodegeneration research program of Amsterdam Neuroscience. Alzheimer Center Amsterdam is supported by Stichting Alzheimer Nederland and Stichting VUmc fonds. The clinical database structure was developed with funding from Stichting Dioraphte. This work was supported by Stichting Alzheimer Nederland (WE.09-2014-06, WE.05-2010-06); Stichting Dioraphte; Internationale Stichting Alzheimer Onderzoek (#11519); JPND-PERADES (ZonMw 733051022): Centralized Facility for Sequence to Phenotype analyses (ZonMW 9111025); Netherlands Consortium for Healthy Aging (NCHA 050-060-810); Biobanking and Biomolecular Research Infrastructure Netherlands (BBMRI-NL CP2010-41); Netherlands Genomics Initiative (NGI)/NWO. This study is further supported by ABOARD, a public-private partnership receiving funding from ZonMW (#73305095007) and Health~Holland, Topsector Life Sciences & Health (PPP-allowance; #LSHM20106). This research is performed by using data from the Parelsnoer Institute an initiative of the Dutch Federation of University Medical Centres ([www.parelsnoer.org](https://eur04.safelinks.protection.outlook.com/?url=http%3A%2F%2Fwww.parelsnoer.org%2F&data=04%7C01%7Ch.holstege%40amsterdamumc.nl%7C2ee28d33af494b36326608d92b2b6190%7C68dfab1a11bb4cc6beb528d756984fb6%7C0%7C0%7C637588287575465763%7CUnknown%7CTWFpbGZsb3d8eyJWIjoiMC4wLjAwMDAiLCJQIjoiV2luMzIiLCJBTiI6Ik1haWwiLCJXVCI6Mn0%3D%7C1000&sdata=ZAqfnkm14AfqX5zuY9EmFnvK2UGeZB5WdTWeCYX5kgY%3D&reserved=0)).

##### 3.2.8 100-plus Study

Cohort collection and exome sequencing of the 100-plus Study cohort was supported by Stichting Alzheimer Nederland (WE.09-2014-03); HorstingStuit Foundation, VUmc Foundation, and the Dioraphte Foundation (Project 17020403), Memorabel (ZonMW project number #733050814, #733050512) and Stichting VUmcFonds. Additional support is from ABOARD, a public-private partnership receiving funding from ZonMW (#73305095007) and Health~Holland, Topsector Life Sciences & Health (PPP-allowance; #LSHM20106).

##### 3.2.9 EMIF-AD 90+

The EMIF-AD 90+ Study was funded by the EU/EFPIA Innovative Medicines Initiative Joint Undertaking EMIF grant agreement no. 115372.

##### 3.2.10 CBC: Control Brain Consortium

This work was supported by the UK Dementia Research Institute which receives its funding from DRI Ltd, funded by the UK Medical Research Council, Alzheimer’s Society and Alzheimer’s Research UK, Medical Research Council (award number MR/N026004/1). Wellcome Trust Hardy (award number 202903/Z/16/Z), Dolby Family Fund; National Institute for Health Research University College London Hospitals Biomedical Research Centre; BRCNIHR Biomedical Research Centre at University College London Hospitals NHS Foundation Trust and University College London.

J. Hardy was supported by the Dolby Foundation and the JPND PERADES. J.B. and R.G were supported by the National Institute on Aging of the National Institutes of Health under Award Number R01AG067426. The content is solely the responsibility of the authors and does not necessarily represent the official views of the National Institutes of Health.

##### 3.2.11 PERADES

We thank all individuals who participated in the study. We also want to express our gratitude to the MRC Centre Core Team for the laboratory support and the Advanced Research Computing at Cardiff University (ARCCA) for the computational support. Cardiff University was supported by the Medical Research Council. Cardiff University was also supported by the European Joint Programme for Neurodegenerative Disease, Alzheimer's Research UK, the Welsh Assembly Government, and a donation from the Moondance Charitable Foundation. Cardiff University acknowledges the support of the UK Dementia Research Institute, which receives its funding from UK DRI Ltd, funded by the UK Medical Research Council, Alzheimer's Society and Alzheimer's Research UK. Cambridge University acknowledges support from the MRC. The University of Southampton acknowledges support from the Alzheimer’s Society. ARUK provided support to Nottingham University. Join Dementia Research (JDR) is funded by the Department of Health and delivered by the National Institute for Health Research in partnership with Alzheimer Scotland, Alzheimer's Research UK and Alzheimer's Society. IRCCS Santa Lucia Foundation acknowledges the Italian Ministry of Health for financial support (IMH_RC) of this study. The Centro de Biologia de Molecular Severo Ochoa (CSIS-UAM), CIBERNED, Instituto de Investigacion Sanitaria la Paz, University Hospital La Paz and the Universidad Autonoma de Madrid were supported by grants from the Ministerio de Educacion y Ciencia and the Ministerio de Sanidad y Consumo (Instituto de Salud Carlos III), and an institutional grant of the Fundacion Ramon Areces to the CMBSO. Thanks to I. Sastre and Dr A Martinez-Garcia for DNA preparation, and Drs P Gil and P Coria for their recruitment efforts. Department of Neurology, University Hospital Mutua de Terrassa, Terrassa, Barcelona, Spain was supported by CIBERNED, Centro de Investigacion Biomedica en Red de Enfermedades Neurodegenerativas, Instituto de Salud Carlos III, Madrid Spain and acknowledges Maria A Pastor (Department of Neurology, University of Navarra Medical School and Neuroimaging Laboratory, Center for Applied Medical Research, Pamplona, Spain), Manuel Seijo-Martinez (Department of Neurology, Hospital do Salnes, Pontevedra, Spain), Ramon Rene, Jordi Gascon and Jaume Campdelacreu (Department of Neurology, Hospital de Bellvitage, Barcelona, Spain) for providing DNA samples. Hospital de la Sant Pau, Universitat Autonoma de Spain acknowledges support from the Spanish Ministry of Economy and Competitiveness (grant number PI12/01311), and from Generalitat de Catalunya (2014SGR-235). The Santa Lucia Foundation and the Fondazione Ca’ Granda IRCCS Ospedale Policlinico, Italy, acknowledge the Italian Ministry of Health (grant RC 10.11.12.13/A)

##### 3.2.12 StEP-AD cohort

The Stanford Extreme Phenotypes in Alzheimer’s Disease (StEP AD) Study is funded by the National Institutes of Health: R01AG060747 and AG047366

##### 3.2.13 Knight-ADRC

NIH P50 AG05681, P01 AG03991, P01 AG026276, NIA U01 AG058922;

##### 3.2.14 UCSF/NYGC/UAB

Funding for genomes sequenced at HudsonAlpha was generously provided by the Daniel Foundation of Alabama and donors to the HudsonAlpha Foundation Memory and Mobility Fund.

##### 3.2.15 UCL-DRC EOAD

This work was supported by the Medical Research Council (UK), the Biomedical Research Centre at University College London Hospitals NHS Foundation Trust and charitable donations to the UCL Dementia Research Centre.

##### 3.2.16 ADSP

The Alzheimer’s Disease Sequencing Project (ADSP) is comprised of two Alzheimer’s Disease (AD) genetics consortia and three National Human Genome Research Institute (NHGRI) funded Large Scale Sequencing and Analysis Centers (LSAC). The two AD genetics consortia are the Alzheimer’s Disease Genetics Consortium (ADGC) funded by NIA (U01 AG032984), and the Cohorts for Heart and Aging Research in Genomic Epidemiology (CHARGE) funded by NIA (R01 AG033193), the National Heart, Lung, and Blood Institute (NHLBI), other National Institute of Health (NIH) institutes and other foreign governmental and non-governmental organizations. The Discovery Phase analysis of sequence data is supported through UF1AG047133 (to Drs. Schellenberg, Farrer, Pericak-Vance, Mayeux, and Haines); U01AG049505 to Dr. Seshadri; U01AG049506 to Dr. Boerwinkle; U01AG049507 to Dr. Wijsman; and U01AG049508 to Dr. Goate and the Discovery Extension Phase analysis is supported through U01AG052411 to Dr. Goate, U01AG052410 to Dr. Pericak-Vance and U01 AG052409 to Drs. Seshadri and Fornage, U54 AG052427 to Drs. Schellenberg and Wang, and R01 AG054060 to Dr Naj. The ADGC cohorts include: Adult Changes in Thought (ACT) (UO1 AG006781, UO1 HG004610, UO1 HG006375, U01 HG008657), the Alzheimer’s Disease Centers (ADC) ( P30 AG019610, P30 AG013846, P50 AG008702, P50 AG025688, P50 AG047266, P30 AG010133, P50 AG005146, P50 AG005134, P50 AG016574, P50 AG005138, P30 AG008051, P30 AG013854, P30 AG008017, P30 AG010161, P50 AG047366, P30 AG010129, P50 AG016573, P50 AG016570, P50 AG005131, P50 AG023501, P30 AG035982, P30 AG028383, P30 AG010124, P50 AG005133, P50 AG005142, P30 AG012300, P50 AG005136, P50 AG033514, P50 AG005681, and P50 AG047270), the Chicago Health and Aging Project (CHAP) (R01 AG11101, RC4 AG039085, K23 AG030944), Indianapolis Ibadan (R01 AG009956, P30 AG010133), the Memory and Aging Project (MAP) ( R01 AG17917), Mayo Clinic (MAYO) (R01 AG032990, U01 AG046139, R01 NS080820, RF1 AG051504, P50 AG016574), Mayo Parkinson’s Disease controls (NS039764, NS071674, 5RC2HG005605), University of Miami (R01 AG027944, R01 AG028786, R01 AG019085, IIRG09133827, A2011048), the Multi-Institutional Research in Alzheimer’s Genetic Epidemiology Study (MIRAGE) (R01 AG09029, R01 AG025259), the National Cell Repository for Alzheimer’s Disease (NCRAD) (U24 AG21886), the National Institute on Aging Late Onset Alzheimer's Disease Family Study (NIA- LOAD) (R01 AG041797), the Religious Orders Study (ROS) (P30 AG10161, R01 AG15819), the Texas Alzheimer’s Research and Care Consortium (TARCC) (funded by the Darrell K Royal Texas Alzheimer's Initiative), Vanderbilt University/Case Western Reserve University (VAN/CWRU) (R01 AG019757, R01 AG021547, R01 AG027944, R01 AG028786, P01 NS026630, and Alzheimer’s Association), the Washington Heights-Inwood Columbia Aging Project (WHICAP) (RF1 AG054023), the University of Washington Families (VA Research Merit Grant, NIA: P50AG005136, R01AG041797, NINDS: R01NS069719), the Columbia University HispanicEstudio Familiar de Influencia Genetica de Alzheimer (EFIGA) (RF1 AG015473), the University of Toronto (UT) (funded by Wellcome Trust, Medical Research Council, Canadian Institutes of Health Research), and Genetic Differences (GD) (R01 AG007584). The CHARGE cohorts are supported in part by National Heart, Lung, and Blood Institute (NHLBI) infrastructure grant HL105756 (Psaty), RC2HL102419 (Boerwinkle) and the neurology working group is supported by the National Institute on Aging (NIA) R01 grant AG033193.

This work was also supported by National Institute on Aging grants R01 AG048927 to Dr. Farrer, RF1 AG054080 to Dr. Beecham, U24 AG056270 to Dr. Mayeux, RF1 AG057519 to Dr. Farrer, U01 AG062602 to Dr. Farrer, R01 AG067501 to Dr. Mayeux, and U19 AG068753 to Dr. Farrer.

The CHARGE cohorts participating in the ADSP include the following: Austrian Stroke Prevention Study (ASPS), ASPS-Family study, and the Prospective Dementia Registry-Austria (ASPS/PRODEM-Aus), the Atherosclerosis Risk in Communities (ARIC) Study, the Cardiovascular Health Study (CHS), the Erasmus Rucphen Family Study (ERF), the Framingham Heart Study (FHS), and the Rotterdam Study (RS). ASPS is funded by the Austrian Science Fond (FWF) grant number P20545-P05 and P13180 and the Medical University of Graz. The ASPS-Fam is funded by the Austrian Science Fund (FWF) project I904),the EU Joint Programme - Neurodegenerative Disease Research (JPND) in frame of the BRIDGET project (Austria, Ministry of Science) and the Medical University of Graz and the Steiermärkische Krankenanstalten Gesellschaft. PRODEM-Austria is supported by the Austrian Research Promotion agency (FFG) (Project No. 827462) and by the Austrian National Bank (Anniversary Fund, project 15435. ARIC research is carried out as a collaborative study supported by NHLBI contracts (HHSN268201100005C, HHSN268201100006C, HHSN268201100007C, HHSN268201100008C, HHSN268201100009C, HHSN268201100010C, HHSN268201100011C, and HHSN268201100012C). Neurocognitive data in ARIC is collected by U01 2U01HL096812, 2U01HL096814, 2U01HL096899, 2U01HL096902, 2U01HL096917 from the NIH (NHLBI, NINDS, NIA and NIDCD), and with previous brain MRI examinations funded by R01-HL70825 from the NHLBI. CHS research was supported by contracts HHSN268201200036C, HHSN268200800007C, N01HC55222, N01HC85079, N01HC85080, N01HC85081, N01HC85082, N01HC85083, N01HC85086, and grants U01HL080295 and U01HL130114 from the NHLBI with additional contribution from the National Institute of Neurological Disorders and Stroke (NINDS). Additional support was provided by R01AG023629, R01AG15928, and R01AG20098 from the NIA. FHS research is supported by NHLBI contracts N01-HC-25195 and HHSN268201500001I. This study was also supported by additional grants from the NIA (R01s AG054076, AG049607 and AG033040 and NINDS (R01 NS017950). The ERF study as a part of EUROSPAN (European Special Populations Research Network) was supported by European Commission FP6 STRP grant number 018947 (LSHG-CT-2006-01947) and also received funding from the European Community's Seventh Framework Programme (FP7/2007-2013)/grant agreement HEALTH-F4- 2007-201413 by the European Commission under the programme "Quality of Life and Management of the Living Resources" of 5th Framework Programme (no. QLG2-CT-2002- 01254). High-throughput analysis of the ERF data was supported by a joint grant from the Netherlands Organization for Scientific Research and the Russian Foundation for Basic Research (NWO-RFBR 047.017.043). The Rotterdam Study is funded by Erasmus Medical Center and Erasmus University, Rotterdam, the Netherlands Organization for Health Research and Development (ZonMw), the Research Institute for Diseases in the Elderly (RIDE), the Ministry of Education, Culture and Science, the Ministry for Health, Welfare and Sports, the European Commission (DG XII), and the municipality of Rotterdam. Genetic data sets are also supported by the Netherlands Organization of Scientific Research NWO Investments (175.010.2005.011, 911-03-012), the Genetic Laboratory of the Department of Internal Medicine, Erasmus MC, the Research Institute for Diseases in the Elderly (014-93-015; RIDE2), and the Netherlands Genomics Initiative (NGI)/Netherlands Organization for Scientific Research (NWO) Netherlands Consortium for Healthy Aging (NCHA), project 050-060-810. All studies are grateful to their participants, faculty and staff. The content of these manuscripts is solely the responsibility of the authors and does not necessarily represent the official views of the National Institutes of Health or the U.S. Department of Health and Human Services.

The four LSACs are: the Human Genome Sequencing Center at the Baylor College of Medicine (U54 HG003273), the Broad Institute Genome Center (U54HG003067), The American Genome Center at the Uniformed Services University of the Health Sciences (U01AG057659), and the Washington University Genome Institute (U54HG003079).
 
Biological samples and associated phenotypic data used in primary data analyses were stored at Study Investigators institutions, and at the National Cell Repository for Alzheimer’s Disease (NCRAD, U24AG021886) at Indiana University funded by NIA. Associated Phenotypic Data used in primary and secondary data analyses were provided by Study Investigators, the NIA funded Alzheimer’s Disease Centers (ADCs), and the National Alzheimer’s Coordinating Center (NACC, U01AG016976) and the National Institute on Aging Genetics of Alzheimer’s Disease Data Storage Site (NIAGADS, U24AG041689) at the University of Pennsylvania, funded by NIA This research was supported in part by the Intramural Research Program of the National Institutes of health, National Library of Medicine. Contributors to the Genetic Analysis Data included Study Investigators on projects that were individually funded by NIA, and other NIH institutes, and by private U.S. organizations, or foreign governmental or nongovernmental organizations.

Data collection and sharing for this project was funded by the Alzheimer's Disease Neuroimaging Initiative (ADNI) (National Institutes of Health Grant U01 AG024904) and DOD ADNI (Department of Defense award number W81XWH-12-2-0012). ADNI is funded by the National Institute on Aging, the National Institute of Biomedical Imaging and Bioengineering, and through generous contributions from the following: AbbVie, Alzheimer’s Association; Alzheimer’s Drug Discovery Foundation; Araclon Biotech; BioClinica, Inc.; Biogen; Bristol-Myers Squibb Company; CereSpir, Inc.; Cogstate; Eisai Inc.; Elan Pharmaceuticals, Inc.; Eli Lilly and Company; EuroImmun; F. Hoffmann-La Roche Ltd and its affiliated company Genentech, Inc.; Fujirebio; GE Healthcare; IXICO Ltd.; Janssen Alzheimer Immunotherapy Research & Development, LLC.; Johnson & Johnson Pharmaceutical Research & Development LLC.; Lumosity; Lundbeck; Merck & Co., Inc.; Meso Scale Diagnostics, LLC.; NeuroRx Research; Neurotrack Technologies; Novartis Pharmaceuticals Corporation; Pfizer Inc.; Piramal Imaging; Servier; Takeda Pharmaceutical Company; and Transition Therapeutics. The Canadian Institutes of Health Research is providing funds to support ADNI clinical sites in Canada. Private sector contributions are facilitated by the Foundation for the National Institutes of Health (www.fnih.org). The grantee organization is the Northern California Institute for Research and Education, and the study is coordinated by the Alzheimer’s Therapeutic Research Institute at the University of Southern California. ADNI data are disseminated by the Laboratory for Neuro Imaging at the University of Southern California.

#### 3.3 Supplementary Authors

Investigators of several cohorts contributed to samples analyzed in this work, but did not participate in analysis or writing of this report:

##### 3.3.1 PERADES Cohort:

Keeley Brookes^1^, Tamar Guetta-Baranes^2^, Elisa Toppi^3^, Francesca Salani^3^, Marina Arcaro^4^, Chiara Fenoglio^5^, Roberta Cecchetti^6^, Elio Scarpini^7^, Sandro Sorbi^8^, Monica Diez-Fairen^9^, Ignacio Alvarez^9^, Miquel Aguilar^9^, MRC: Simon Lovestone^10^, John Powell^11^, Carol Brayne^12^, David Rubinsztein^12^, Nandini Badarinarayan^13^, Eloy Rodriguez-Rodriguez^14^, Carmen Lage^14^, Sara Lopez-Garcia^14^, Emanuele Costantini^15^, Michela Orsini^15^, Francesco Panza^16^, Nerisa Banaj^17^, Federica Piras^17^ and Daniela Vecchio^17^

**(1)**Nottingham Trent; **(2 )**Human Genetics. UoN; (**3**) IRCCS Fondazione Santa Lucia, Department of Clinical and Behavioral Neurology, Experimental Neuro-psychobiology Lab Via Ardeatina, 306, I-00179 Roma, Italy; **(4)** Fondazione IRCCS Ca' Granda, Ospedale Policlinico; **(5)** University of Milan, Dino Ferrari Center, Milan, Italy; 6Institute of Gerontology and Geriatrics, Department of Medicine and Surgery, University of Perugia, Italy; **(7)** University of Milan, Centro Dino Ferrari, CRC Molecular basis of Neuro-Psycho-Geriatrics diseases, Milan, Italy; **(8)** Department of Neuroscience, Psychology, Drug Research and Child Health , University of Florence, Italy; **(9)** Memory Disorders Unit, Department of Neurology, Hospital Universitari Mutua de Terrassa, Terrassa, Barcelona, Spain; **(10)** Department of Psychiatry, Medical Sciences Division, University of Oxford, Oxford, UK.; **(11)** Kings College London, Institute of Psychiatry, Department of Neuroscience, De Crespigny Park, Denmark Hill, London, UK; **(12)** Institute of Public Health, University of Cambridge, Cambridge, UK.; (**13)** Division of Psychological Medicine and Clinical Neuroscience, School of Medicine, Cardiff University, Cardiff, UK; **(14)** Neurology Service and Centro de Investigación en Red de Enfermedades Neurodegenerativas (CIBERNED), Marques de Valdecilla University Hospital (University of Cantabria and IDIVAL), Santander, Spain **(15)** Department of Neuroscience, Catholic University of Sacred Heart, Fondazione Policlinico Universitario A. Gemelli IRCCS, Rome, Italy **(16)** National Institute of Gastroenterology and Research Hospital IRCCS “S. De Bellis” Castellana Grotte, Bari Italy **(17)** Laboratory of Neuropsychiatry,IRCCS Santa Lucia Foundation, Rome, Italy

##### 3.3.2 StEP AD investigators

Clifton L. Dalgard, PhD^1^, William J. Jagust, MD^2^, Sterling C. Johnson, PhD	^3^, David A. Wolk, MD^4^, Joel H. Kramer, PsyD^5^, Bradford C. Dickerson, MD^6^, David A. Bennett, MD^7^,

Sofiya Milman, MD^8^, Bruno Dubois, MD, PhD^9^, Ruth O’Hara, PhD^10^, Sherry A. Beaudreau, PhD^11^

^1^Uniformed Services University of the Health Sciences;^2^University of California, Berkeley; ^3^University of Wisconsin; ^4^University of Pennsylvania; ^5^University of California, San Francisco; ^6^Harvard University; ^7^Rush University; ^8^Albert Einstein College of Medicine; ^9^Brain and Spine Institute (ICM), France; ^10^Stanford University

^11^VA Palo Alto Health Care System

##### 3.3.3 Knight ADRC investigators

Achal Neupane^1,3,4^, John P Budde^1,3,4^, Fengxian Wang^1,3,4^, Joanne Norton^1,3,4^, Gen Gentsch^1,3,4^, John C Morris^2,3^

Departments of ^1^Psychiatry, ^2^Neurology, ^3^Hope Center for Neurological Disorders, ^4^NeuroGenomics and Informatics Center, Washington University School of Medicine, St. Louis, Missouri, USA.

##### 3.3.4 ADNI database

A subset of the data used in this article was obtained from the Alzheimer’s Disease Neuroimaging Initiative (ADNI) database (adni.loni.usc.edu). A complete listing of ADNI

investigators can be found at:

http://adni.loni.usc.edu/wp-content/uploads/how_to_apply/ADNI_Acknowledgement_List.pdf
